# Impact of viral epidemic outbreaks on mental health of healthcare workers: a rapid systematic review

**DOI:** 10.1101/2020.04.02.20048892

**Authors:** Ignacio Ricci-Cabello, Jose F. Meneses-Echavez, Maria Jesús Serrano-Ripoll, David Fraile-Navarro, Maria Antònia Fiol de Roque, Guadalupe Pastor Moreno, Adoración Castro, Isabel Ruiz-Pérez, Rocío Zamanillo Campos, Daniela Gonçalves-Bradley

## Abstract

**Objectives:** To examine the impact of providing healthcare during or after health emergencies caused by viral epidemic outbreaks on healthcare workers′(HCWs) mental health, and to assess the available evidence base regarding interventions to reduce such impact.

**Design:** Systematic rapid review and meta-analysis.

**Data sources:** MEDLINE, Embase, and PsycINFO, searched up to 23 March 2020.

**Method:** We selected observational and experimental studies examining the impact on mental health of epidemic outbreaks on HCWs. One reviewer screened titles and abstracts, and two reviewers independently reviewed full texts. We extracted study characteristics, symptoms, prevalence of mental health problems, risk factors, mental health interventions, and its impact. We assessed risk of bias for each individual study and used GRADE to ascertain the certainty of the evidence. We conducted a narrative and tabulated synthesis of the results. We pooled data using random-effects meta-analyses to estimate the prevalence of specific mental health problems.

**Results:** We included 61 studies (56 examining impact on mental health and five about interventions to reduce such impact). Most were conducted in Asia (59%), in the hospital setting (79%), and examined the impact of the SARS epidemic (69%). The pooled prevalence was higher for anxiety (45%, 95% CI 21 to 69%; 6 studies, 3,373 participants), followed by depression (38%, 95% CI 15 to 60%; 7 studies, 3,636 participants), acute stress disorder (31%, 95% CI 0 to 82%, 3 studies, 2,587 participants), burnout (29%, 95% CI 25 to 32%; 3 studies; 1,168 participants), and post-traumatic stress disorder (19%, 95% CI 11 to 26%, 10 studies, 3,121 participants). Based on 37 studies, we identified factors associated with the likelihood of developing those problems, including sociodemographic (younger age and female gender), social (lack of social support, social rejection or isolation, stigmatization), and occupational (working in a high risk environment (frontline staff), specific occupational roles (e.g., nurse), and lower levels of specialised training, preparedness and job experience) factors. Five studies reported interventions for frontline HCW, two of which were educational and aimed to prevent mental health problems by increasing HCWs′ resilience. These interventions increased confidence in support and training, pandemic self-efficacy, and interpersonal problems solving (very low certainty). One multifaceted intervention implemented training and organisational changes) targeted at hospital nurses during the SARS epidemic, reporting improvements in anxiety, depression, and sleep quality (very low certainty). The two remaining interventions, which were multifaceted and based on psychotherapy provision, did not assess their impact.

**Conclusion:** The prevalence of anxiety, depression, acute and post-traumatic stress disorder, and burnout, was high both during and after the outbreaks. These problems not only have a long-lasting effect on the mental health of HCWs, but also hinder the urgent response to the current COVID-19 pandemic, by jeopardising attention and decision-making. Governments and healthcare authorities should take urgent actions to protect the mental health of HCWs. In light of the limited evidence regarding the impact of interventions to tackle mental health problems in HCWs, the risk factors identified in this study, more so when they are modifiable, represent important targets for future interventions.

**SUMARY BOX:** *What is already known on this topic?:*

- Previous studies showed that healthcare workers involved providing frontline care during viral epidemic outbreaks are at high risk of developing mental health problems.
- Given the current COVID-19 pandemic, there is an urgent need to synthesize the evidence regarding the impact of viral epidemic outbreaks on mental health of healthcare workers.

*What does this study add?:*

- This timely systematic rapid review offers for the first time pooled estimations of the prevalence of the most common mental health problems experienced by HCWs during and after viral epidemic outbreaks, namely anxiety (45%), depression (38%), and acute stress disorder (31%), among others.
- Our study also identifies a broad number of factors associated with these conditions, including sociodemographic factors such as younger age and female gender, social factors such as lack of social support, social rejection or isolation, stigmatization, and occupational factors such as working in a high risk environment, specific occupational roles, and having lower levels of specialised training, preparedness and job experience.
- Our study shows that, although educational and multifaceted interventions might mitigate the development of mental health problems, the certainty on the evidence is very low - therefore indicating that further high quality research is urgently needed to inform evidence-based policies for viral pandemics.

## INTRODUCTION

Infectious disease outbreaks are relatively common,^1^ often prompting an international response involving thousands of healthcare workers (HCWs).^2^ Providing frontline healthcare during infectious outbreaks increases the risk of HCWs developing mental health problems, both short and long-term.^3^ It has been suggested that specific occupational factors are associated with psychological outcomes of HCWs during an infectious disease outbreak.^2^ Working in a high-risk environment, adhering to quarantine, job-related stress, and belonging to a specific cadre were all considered to aggravate psychological outcomes. Perceived safety, namely through access to protective equipment, and specialised training, mitigated those outcomes.^2^

During December 2019 a new infectious disease outbreak was reported in Wuhan, Hubei province, China,^4^ which was named COVID-19.^5^The World Health Organization (WHO) declared COVID-19 a pandemic by March 11th 2020, and by 30 March 2020 it had spread to most countries and territories, with more than 693,000 known cases and a death toll of over 33,000 people.^6^ Early anecdotal evidence from Wuhan showed how this unprecedented situation impacted the mental health of frontline HCWs, who reported mental problems such as anxiety, depressive symptoms, anger, and fear.^7^ These problems cannot only have a long-lasting effect on the mental health of HCWs,^3^ but also hinder the urgent response to COVID-19, by jeopardising attention and decision-making.^7^ Tackling the mental health of HCWs during this pandemic is essential, and will strengthen healthcare systems’ capacity.^8^

Previous systematic reviews have explored social and occupational factors associated with psychological outcomes in HCW during an infectious disease outbreak,^2^ and their perceptions of risk and use of coping strategies towards emerging respiratory infectious diseases.^9^ However, to date, the impact of viral disease outbreaks on specific mental health problems and the effectiveness of interventions to ameliorate such impact have not been systematically reported. The aim of this rapid systematic literature review is twofold: i) to examine the impact of health emergencies caused by a viral pandemic or epidemic on HCWs mental health; and ii) to assess the effectiveness of interventions to reduce such impact.

## METHODS

We conducted a rapid systematic review following WHO guidelines^10^ and Cochrane’s recommendations for Rapid Reviews in response to COVID-19.^11^ We followed the Preferred Reporting Items for Systematic Reviews and Meta-Analyses (PRISMA) guidelines for planning, conducting and reporting this study.^12^

### Data Sources and Searches

We designed specific search strategies for biomedical databases (MEDLINE/Ovid, EMBASE/Elsevier, and PsycInfo/EBSCO), combining MeSH terms and free-text keywords (Online Appendix 1). We searched databases from inception to 23rd March 2020, and checked the list of included studies of relevant systematic reviews.^9 13 14^ We used EndNote X8™ to create a bibliographical database, and Rayyan to screen relevant records.^15^

### Selection criteria

We included empirical studies examining the impact on mental health of epidemic outbreaks on HCWs, and studies about interventions to reduce such impact. We included observational (cross-sectional, case-control, and cohort studies), and experimental studies (non-controlled before-after studies, controlled before-after studies, non-randomised controlled trials, and randomised controlled trials).

We included studies on any type of health emergency caused by a viral epidemic or pandemic, and examining its impact on HCWs mental health during or after the crisis. For intervention studies, we included also those that examined interventions to protect mental health of healthcare workers prior, during or after the outbreak onset. All types of settings and healthcare professionals were accepted for inclusion. We included studies measuring any type of mental health problem or psychiatric morbidity. We excluded narrative reviews, thesis, editorials, protocols, letters to the editor, and studies published in languages other than English, Spanish or Portuguese.

### Study Selection

One reviewer (of IRC, MJSR, MAFR, RZC, DGB) screened the retrieved references at title and abstract against the selection criteria. Two reviewers (of those aforementioned) independently and blinded against the others’ judgements assessed full-text eligibility. We solved disagreements by consensus or by involving a third reviewer, if needed.

### Data Extraction and Quality Assessment

We used structured forms to extract relevant data, such as country, health emergency, setting, population, epidemiological design, number of participants, mental health conditions, clinical outcomes and their measurement tools, and main study results. For observational studies addressing the impact of health emergencies on HCWs mental health, we extracted the prevalence rate of the mental conditions examined in terms of the number of professionals suffering the condition (numerator) out of the total number of study participants (denominator). If available, we extracted information about the risk factors. For intervention studies (i.e., randomised and non-randomised trials), we extracted data about the characteristics of the intervention as well as that reported also for observational studies. We assessed the risk of bias of observational studies (i.e., cross-sectional, case-control, and cohort studies) by using the set of tools developed by Evidence Partners (McMaster University) ^16^; whereas ROBINS I ^17^ was applied to uncontrolled trials, and AMSTAR ^18^for systematic reviews.

One reviewer (of MJSR, MAFR, AC, DF, JM, GP, RZ) extracted all the data and assessed the risk of bias, while a second reviewer cross-checked the information for accuracy and completeness.

### Data Synthesis and Analysis

We conducted a narrative and tabulated synthesis of the results, classifying the studies according to the type of study (i.e., impact of infectious disease outbreaks on HCWs mental health, or interventions to reduce such impact), and timing (i.e., before, during, or after the outbreak). We adapted a taxonomy proposed in a previous study ^14^ to classify risk factors as social, occupational and sociodemographic.

For studies about the impact of outbreaks on mental health, we conducted random-effects meta-analyses to estimate the prevalence and 95% confidence interval (CI) of each type of mental health condition, using the STATA command “metaprop”. We conducted subgroup analyses to explore potential differences in the prevalence of mental health disorders during vs. after the outbreak. Heterogeneity was quantified by the *I*^2^ statistic, where *I*^2^>50% was deemed as substantial heterogeneity.^19^ Publication bias was examined with funnel plots and presence of asymmetry tested with Begg^20^ and Egger tests.^21^ We used Stata, version 12.0 to conduct meta-analyses.

### GRADE and ‘Summary of findings’ tables

We used the GRADE approach^22^ to assess the quality of evidence related to the outcomes included in this rapid review. We used GRADEpro 2011 ^23^ software to create ‘Summary of findings’ tables. For assessments of the overall quality of evidence for each outcome that included pooled data, we downgraded the evidence from ‘high quality’ by one level for serious, or by two levels for very serious, study limitations (risk of bias), indirectness of evidence, inconsistency, imprecision of effect estimates, or potential publication bias.^22^

### Patient and public involvement

We have invited HCW frontline to promote the dissemination of these results, alongside members of the author team who are also frontline HCWs.

## RESULTS

### Search results

The search resulted in a total of 2,317 records. After 143 duplicates were removed, 2,174 records remained to be screened. We excluded 2,042 records based on title and abstract screening. We assessed full-text 132 articles in full-text, of which we excluded 74. After including three additional studies identified from manual searches, sixty-one published studies met the inclusion criteria for this systematic rapid review.^13 24-83^ Figure 1 illustrates the selection process of the included studies. Online Appendix 2 presents the excluded studies.

**Figure 1.**
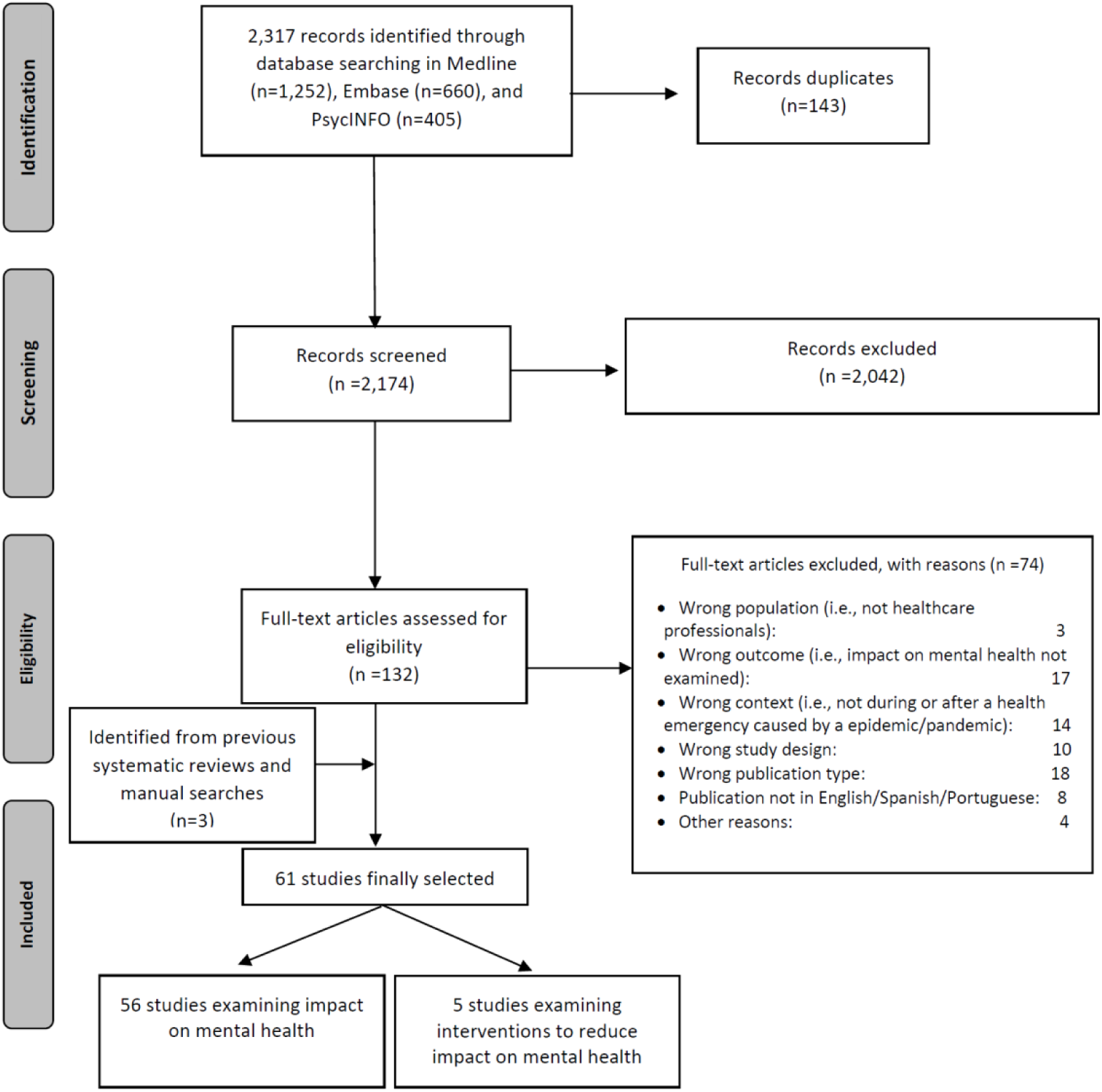
PRISMA flow diagram

### Characteristics of the studies

This systematic rapid review included 38,415 participants (total). Most of the studies (59%) were conducted in Asian countries, including China (30%), South Korea (18%), Taiwan (15%) or Singapore (12%). The mean number of participants was 612 (range 26 to 10,511). More than two-thirds (69%) examined the impact of Severe acute respiratory syndrome (SARS epidemic), followed by Middle East respiratory syndrome coronavirus (MERS-CoV) (11%). Three studies examined the impact of COVID-19.^46 53 83^ Most studies were conducted during or after the infectious outbreak (92%). Around three quarters took place in the hospital setting. General HCWs was the most common group (70%), followed by nurses (20%) and physicians (10%). Post-traumatic stress disorder (PTSD) (38%), anxiety (36%), depression (28%) and stress/distress symptoms (28%) were the mental health conditions most frequently examined. The majority followed a cross-sectional design (82%). The characteristics of the included studies are summarised in Table 1.

**Table 1.** Features of the studies selected (N=61)

|  | N | % |
| --- | --- | --- |
| **Year of the study publication** |  |  |
| 2001-2005 | 22 | 36 |
| 2006-2010 | 19 | 31 |
| 2011-2015 | 5 | 8 |
| 2016-2020 | 15 | 25 |
| **Epidemiologic design** |  |  |
| Cross-sectional | 50 | 82 |
| Cohort study | 7 | 11 |
| Quasi-experimental | 2 | 3 |
| Case-control | 1 | 2 |
| Systematic literature reviews | 1 | 2 |
| **Number of participants \*** | 612 | (26 - 10,511) |
| **Mental health problems.↓** |  |  |
| Post-traumatic stress disorder | 23 | 38 |
| Anxiety | 22 | 36 |
| Depression | 17 | 28 |
| Stress/distress | 17 | 28 |
| Burnout | 8 | 13 |
| Acute stress disorder | 7 | 11 |
| Mental health status (overall assessment) | 20 | 33 |
| Others | 15 | 25 |
| **Country.↓** |  |  |
| China | 18 | 30 |
| Canada | 12 | 20 |
| South Korea | 11 | 18 |
| Taiwan | 9 | 15 |
| Singapore | 7 | 12 |
| Saudi Arabia | 3 | 5 |
| Others | 11 | 18 |
| **Study timing** |  |  |
| After outbreak | 28 | 46 |
| During outbreak | 28 | 46 |
| Both during and after outbreak | 2 | 3 |
| Prior, during and after outbreak | 2 | 3 |
| Prior outbreak onset | 1 | 2 |
| **Type of health emergency** |  |  |
| SARS | 42 | 69 |
| MERS-COV | 7 | 11 |
| H1N1 influenza virus | 4 | 7 |
| Ebola | 4 | 7 |
| COVID-19 | 3 | 5 |
| H7N9 influenza virus | 1 | 2 |
| **Population** |  |  |
| Health care workers in general | 43 | 70 |
| Nurses | 12 | 20 |
| Doctors | 6 | 10 |
| **Setting** |  |  |
| Hospital | 48 | 79 |
| Healthcare facilities in general | 8 | 13 |
| Primary Care centre | 3 | 5 |
| Non specified | 2 | 3 |

\* mean and range, ↓ percentages exceeding 100% as categories are not mutually exclusive.

### Risk of bias assessment

In general, main risks of bias in the 50 cross-sectional studies were the lack of use of reliable and valid instruments to measure mental health outcomes (high risk of bias in 22% of the studies) and selection bias (12%). The main sources of bias across the seven cohort studies were selection bias (43%) and inadequate follow-up of the cohorts (29%). Main sources of bias of the two uncontrolled before-after studies were bias in selection of participants, and bias in outcome measurement. The case-control and the systematic review identified did not present serious risks of bias. Results of the risk of bias assessment are provided in Online Appendix 3.

### Prevalence of mental health problems in HCWs during and after infectious disease outbreaks

Fifty-six studies examined the mental health problems among frontline HCWs during and/or after an infectious disease outbreak (Online Appendix 4). The great majority of them reported clinically significant mental health symptoms, most frequently PTSD, anxiety, depression, and burnout. For clinically significant symptoms of mental health disorders, the pooled prevalence was higher for anxiety (45%, 95% CI 21 to 69%, I*2* 99.7%; 6 studies, 3,373 participants), followed by depression (38%, 95% CI 15 to 60%, *I2* 99.6%; 7 studies, 3,636 participants), and PTSD (19%, 95% CI 11 to 26%, *I2* 97.6%; 10 studies, 3,121 participants). Three studies reported the prevalence of burnout (29%, 95% CI 25 to 32%; 1,168 participants), and three reported the prevalence of acute stress disorder (31%, 95% CI 0 to 82%, 2,587 participants). Subgroup analyses found little or no differences in prevalence during vs. after the outbreaks (Online Appendix 5). Begg’s and Egger’s tests suggested the absence of publication bias for all the meta-analyses conducted.

### Risk factors for mental health problems in HCWs during and after infectious disease outbreaks

Thirty-seven studies examined a large number of occupational, sociodemographic and social factors associated with the likelihood of developing mental health problems while providing frontline healthcare during an infectious disease outbreak (Online Appendix 4).

The main occupational factors were working in a high risk environment, higher perception of threat and risk, specialised training received, and specific occupational role. Working in a high risk environment was associated with different mental health problems, namely depression,^46^ anxiety,^46 52 60 77^ PTSD,^28 72 75 81 82^ and burnout.^76^ The definition of *high risk environment* varied across studies, but usually included being in direct contact with infected patients, either providing care,^28 77^ or being responsible for cleaning and disinfection.^52^

Likewise, higher perception of threat and risk was also associated with a higher prevalence of a number of different mental health problems, including depression,^55^ anxiety,^25^ and PTSD.^63 72 81^ Lack of specialised training was a risk factor for anxiety,^60 79^ PTSD,^75^ and burnout.^63^

Some of the studies that recruited more than one cadre reported that specific HCWs were at higher risk of developing mental health problems. One study each found that nurses were more likely to develop PTSD ^75^ and burnout,^76^ whereas one study ^26^ reported that resident pulmonologists were at higher risk of burnout.

Other occupational risk factors for PTSD were job stress,^63^ and less job experience,^75^ whereas lower levels of organisational support increased the risk of burnout.^59^

Studies addressing sociodemographic and social risk factors focused on PTSD and burnout. Younger age was a risk factor for both PTSD ^75^ and burnout,^26^ while female gender was associated with higher levels of PTSD in HCWs.^75^ Feelings of social rejection or isolation,^63^ and higher impact of the outbreak on daily life ^72^ increased the likelihood of developing PTSD, whereas lack of family and friends support were associated with burnout.^43^ In addition, stigmatisation,^44^ social rejection,^67^ and lower levels of social support were identified as risk factors for stress.^83^

### Interventions to reduce the mental health impact of viral outbreaks in HCWs

Five studies ^24 33 40 62 70^ described five different interventions to reduce the mental health impact of viral outbreaks in HCWs (Online Appendix). Two studies implemented in Canada evaluated two educational interventions for improving HCWs mental health by increasing resilience.^24 62^ Aiello and colleagues^24^ described an educational intervention targeted to HCWs during the SARS epidemic, which consisted of a face-to-face group training session based on Folkman and Greer’s model of coping.^84^ The session focused on stressors associated with pandemic influenza and on organisational and individual approaches to building resilience and reducing stress. While most participants did not feel prepared to deal confidently with the pandemic before the session (35%), there was a higher proportion of participants who felt better able to cope after the session (76%).

Maunder and colleagues explored the impact of a computer-assisted resilience training to prepare HCWs for a potential pandemic influenza.^62^ The course consisted of modules incorporating different modalities of learning (knowledge-based modules, relaxation skills, and self-assessment modules using questionnaires to characterize interpersonal problem and coping style). The intervention improved confidence in support and training, pandemic self-efficacy and interpersonal problems (p<0.05). We have very low confidence on the evidence of educational interventions for preventing the psychological impact of infectious epidemic outbreaks in HCWs (detailed in Online Appendix 7) due to the study design (uncontrolled before-after studies) and very serious risk of bias with regard to confounding and measurement of outcomes.

Two studies examined two multifaceted interventions combining training and implementation of organizational changes.^33 70^ A study in Taiwan ^33^ evaluated the effects of a multifaceted intervention to prevent depression and anxiety in hospital nurses during the SARS epidemic. The intervention included in-service training, manpower allocation, gathering sufficient protective equipment, and establishment of a mental health team. The authors observed statistically significant improvements in nurses’ anxiety and depression along with sleep quality at two weeks follow-up. Another study described a multifaceted intervention to improve resilience and prevent PTSD in HCWs during the Ebola epidemic in the USA, Philippines, and West Africa.^70^ The intervention, based on the Anticipate, Plan and Deter Responder Risk and Resilience model, included pre-deployment development of an individualized resilience plan and an in-theatre, real-time self-triage system, to allow HCWs to assess and manage the full range of psychological risk and resilience for themselves and their families. The potential effectiveness of this intervention was not studied. Our confidence on the evidence for multifaceted interventions for preventing the psychological impact during infectious epidemic outbreaks in HCWs was very low (Online Appendix 7) due to limitations in the study design (uncontrolled before after studies) and very serious risk of bias (high risk of selection bias and high risk of bias in measurement of outcomes).

Finally, Khee et al. 2004 reported an intervention in 188 hospital nurses in Singapore, consisted in the provision of psychological support during the SARS outbreak.^40^ The intervention, not based on any specific psychotherapeutic model, comprised multiple sessions (75 minutes per session) and was aimed at preserving their mental health. The primary goal of therapy was to externalise all their emotions, and bring support to each other. The effectiveness of this intervention was not studied.

## DISCUSSION

### Summary of findings

In this timely systematic rapid review we synthesized evidence from 61 studies examining the impact on mental health of providing frontline healthcare during infectious disease outbreaks. Results showed that HCWs commonly present high levels of anxiety, depression, PTSD, acute disorder and burnout, both during and after the outbreaks. We identified a broad number of risk factors for these conditions, including sociodemographic factors such as younger age and female gender, and social factors such as lack of social support, social rejection or isolation, stigmatization. Occupational factors entailed working in a high risk environment (frontline staff), specific occupational roles (e.g., nurse), and having lower levels of specialized training, preparedness and job experience. In contrast with the high number of studies examining impact on mental health, there is limited evidence regarding the impact of interventions to reduce mental health problems in this particularly vulnerable population, and overall its certainty is very low, mainly due to study design and serious risk of bias.

### Strengths and limitations of the review

This is a timely and comprehensive rapid review of the current literature on the impact of infectious disease outbreaks on the mental health of HCWs. We examined three relevant areas, namely the prevalence of mental health problems, factors associated with an increased likelihood of developing those problems, and the effects of interventions to improve mental health of HCWs. We followed the highest methodological standards when undertaking the current rapid review,^10^ and we used the GRADE approach to evaluate the certainty of the evidence, in order to facilitate evidence-informed decision making processes. Our review team is also a strength, as it included experts in evidence synthesis, Cochrane authors, members of the GRADE Working Group, physicians, nurses, editors, psychologists, and psychiatrists. There were also some limitations underlying this work. Despite searching three major databases and manually searching references of previously published systematic reviews, we did not examine gray literature; hence, we cannot discard that relevant references may have been missed out. We observed high heterogeneity when pooling data, which could be partially attributed to the high variability across studies in terms of study population (e.g. occupational role), context (e.g. magnitude of the health emergency caused by epidemic) and outcome measures. In light of this, our results should be interpreted with caution.

## Discussion of the main findings

Some of the risk factors associated with mental health problems while providing frontline care during infectious disease outbreaks cannot be modified. In this way, working in a high risk environment increases the risk of developing clinically significant symptoms, namely depression,^46^ anxiety,^46 52 60 77^ PTSD,^28 72 75 81 82^ and burnout.^76^ Likewise, it seems like specific cadres are more likely to report mental health problems, namely PTSD,^75^ and burnout.^26 76^

However this review also identified specific modifiable factors that can be addressed in advance and mitigate the risk brought by the aforementioned factors. Lack of specialized training was associated with anxiety, ^60 79^ PTSD,^75^ and burnout,^63^ and higher perception of threat and risk was associated with depression,^55^ anxiety,^25^ and PTSD ^38 63 72 81^. Long-term institutional preparedness is possible for both factors, through the development and implementation of specialized training that includes infection prevention, diagnostics, patient care, staff, and communication.^85^

Continuous communication between HCWs and managers, including the provision of up-to-date facts about the progression of the outbreak, can convey institutional support,^59^ and promote the acquisition of knowledge and confidence for those HCWs who have less job experience.^75^ Likewise, managers are essential to mitigate feelings of social isolation ^50 63^ and stigmatization, ^44^ especially among those HCWs who have to be quarantined.^13^ The proliferation of online mobile-based technologies will play an essential role in promoting connectedness and decrease the feelings of isolation and stigmatization,^86^ and can also be used for informal contacts between HCWs who are quarantined.

Although limited, evidence from intervention studies indicates that educational interventions have the potential to increase knowledge and resilience, ^24 62^ even when implemented during an outbreak.^33^

### Limitations of available evidence and future research needs

We identified 56 studies reporting on the impact on mental health of providing frontline healthcare during an infectious disease outbreak, however most of the studies did not use validated methods to assess mental health, which limits the generalizability of our findings. Furthermore, only a handful of studies assessed the efficacy of interventions to ameliorate the impact of health emergencies on mental health of HCWs.

It is expected the proliferation of a large volume of studies examining the impact of COVID-19 on HCWs′ mental health during the near future. To make progress in this area, future studies should address these limitations of the available literature. The use of validated measurement tools and more representative sample sizes are warranted in order to strengthen the quality of the evidence in this area. Intervention studies should also adhere to international reporting standards such as CONSORT ^85^ and TIDieR.^87^

## Conclusions

As we demonstrated in our review, the mental health burden for HCWs during pandemics is especially high both during and after the outbreak. Of note for its similarity to the current COVID-19 crisis, are the experiences gained from the previous SARS outbreak. This time, given the size, scale and importance of the current pandemic, these trends could be much worse.

We urge governments, policy makers and relevant stakeholders to monitor and follow these outcomes and conduct scientifically sound interventional research, in order to mitigate mental health impact on HCWs.

The physical health of HCWs is already at stake from the virus, and once we tackle the current pandemic, we will need to heal the healers, not only for the sake of having a prepared and resilient work-force, but as we owe them from the tremendous sacrifices they are doing. If we want to address these concerns and be able to mitigate its impact, we need to act soon.

## Data Availability

No additional data available

## Contributors

IRC, IRP and MJSR had the idea for the study. IRC designed the search strategy. IRC, MJSR, MAFR, RZC, DGB screened abstracts and full texts. MJSR, MAFR, AC, DFN, JM, GP, RZ, DGB acquired data, and assessed risk of bias in studies. IRC did the data analysis. All authors interpreted the data analysis. IRC and DGB wrote the manuscript, with revisions from all authors. The corresponding author attests that all listed authors meet authorship criteria and that no others meeting the criteria have been omitted. IRC is the guarantor.

## Funding

No specific funding for this study. IRC is a recipient of a Miguel Servet Fellowship (project number CP17/00017) funded by the Spanish Government. The funding sources had no role in the design and conduct of the study; collection, management, analysis, and interpretation of the data; preparation, review, or approval of the manuscript; and decision to submit the manuscript for publication.

### Competing interests

All authors have completed the ICMJE uniform disclosure form at www.icmje.org/coi_disclosure.pdf and declare: no support from any organisation for the submitted work; no financial relationships with any organisations that might have an interest in the submitted work in the previous three years; no other relationships or activities that could appear to have influenced the submitted work.

## Ethical approval

Not required

## Data sharing

No additional data available.

## Transparency

The lead author affirms that the manuscript is an honest, accurate, and transparent account of the study being reported; that no important aspects of the study have been omitted; and that any discrepancies from the study as planned (and, if relevant, registered) have been explained.

## Online Appendix 1 Search strategies

Medline (Ovid): 23 March 2020

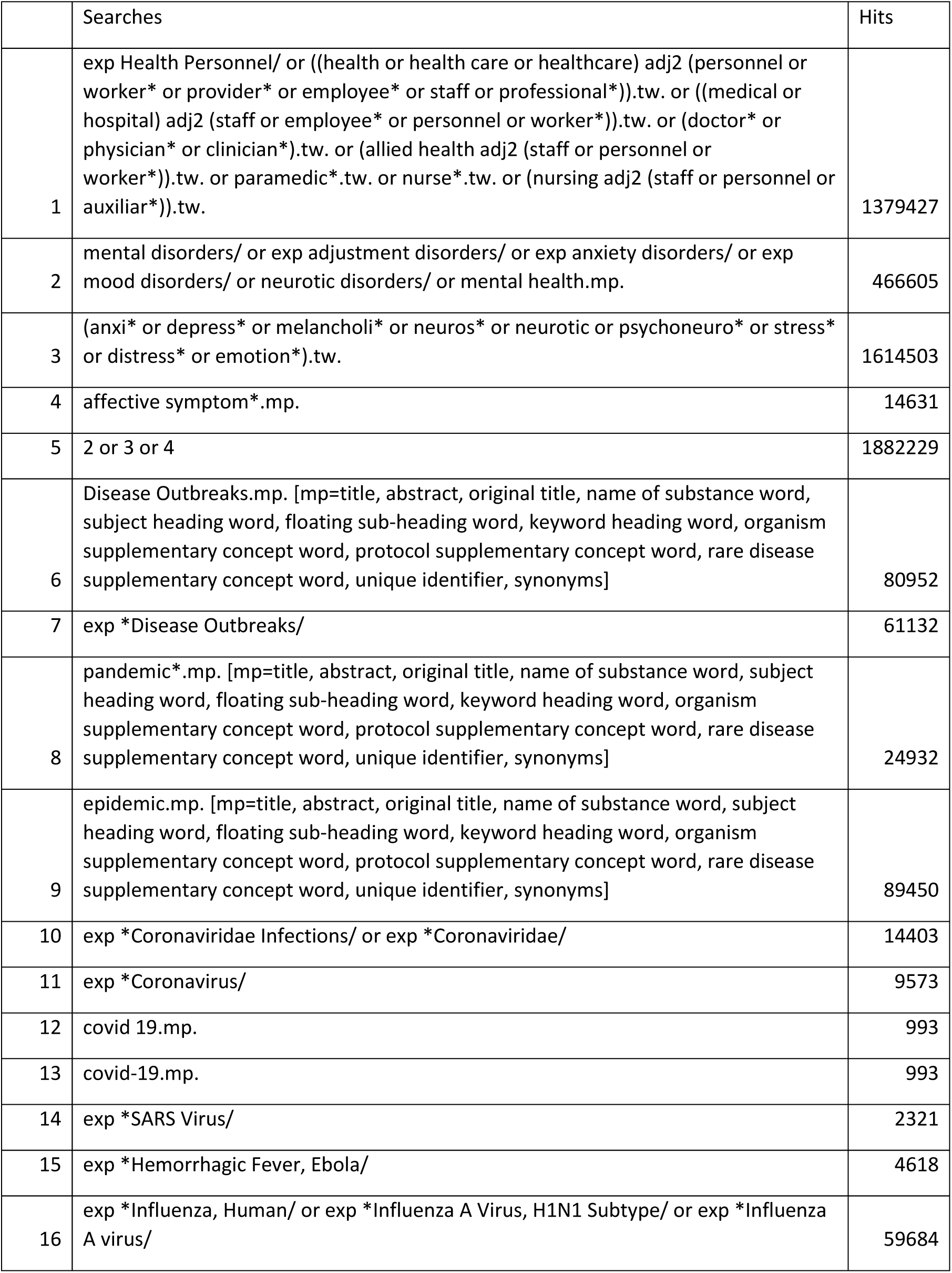
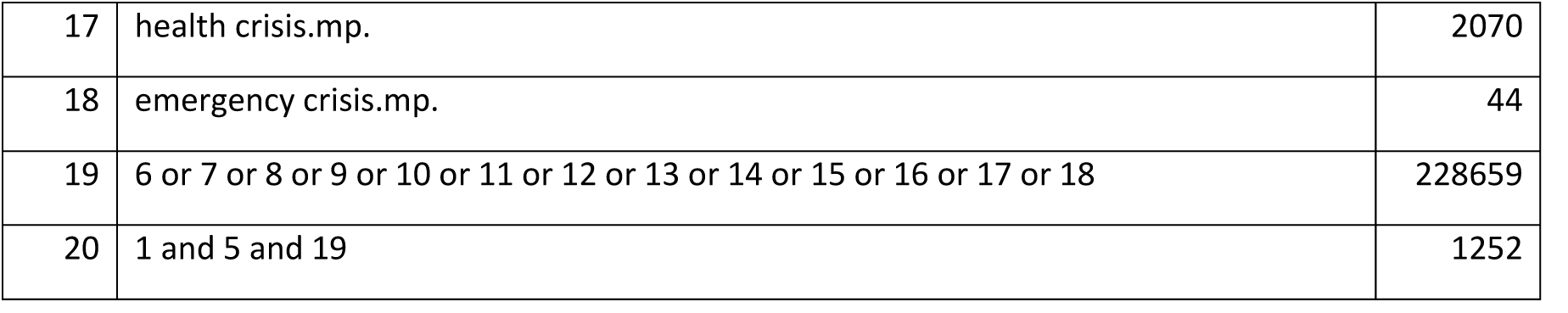

PsycINFO (EBSCO): 23 March 2020

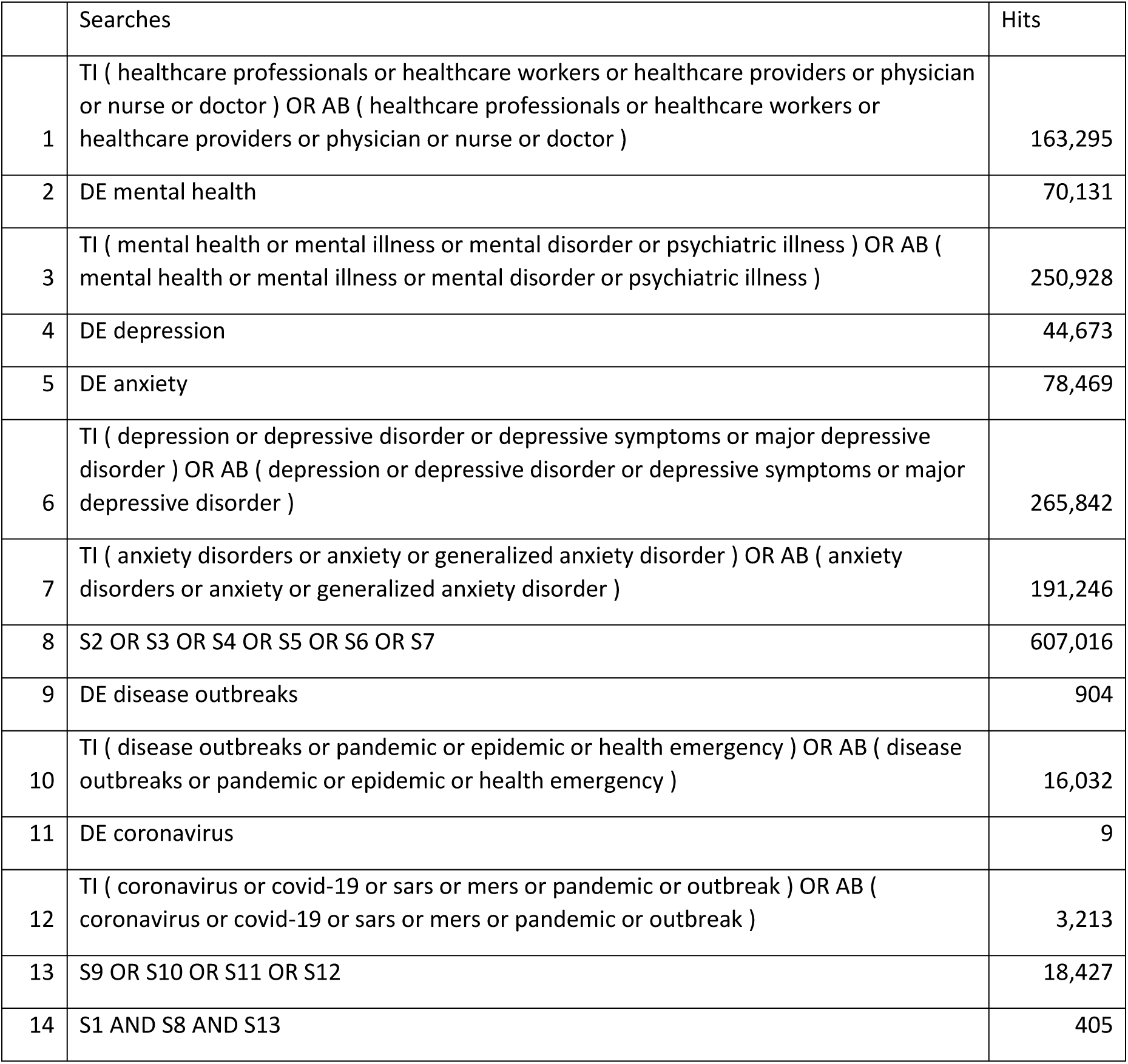

Embase (Elsevier): 23 March 2020

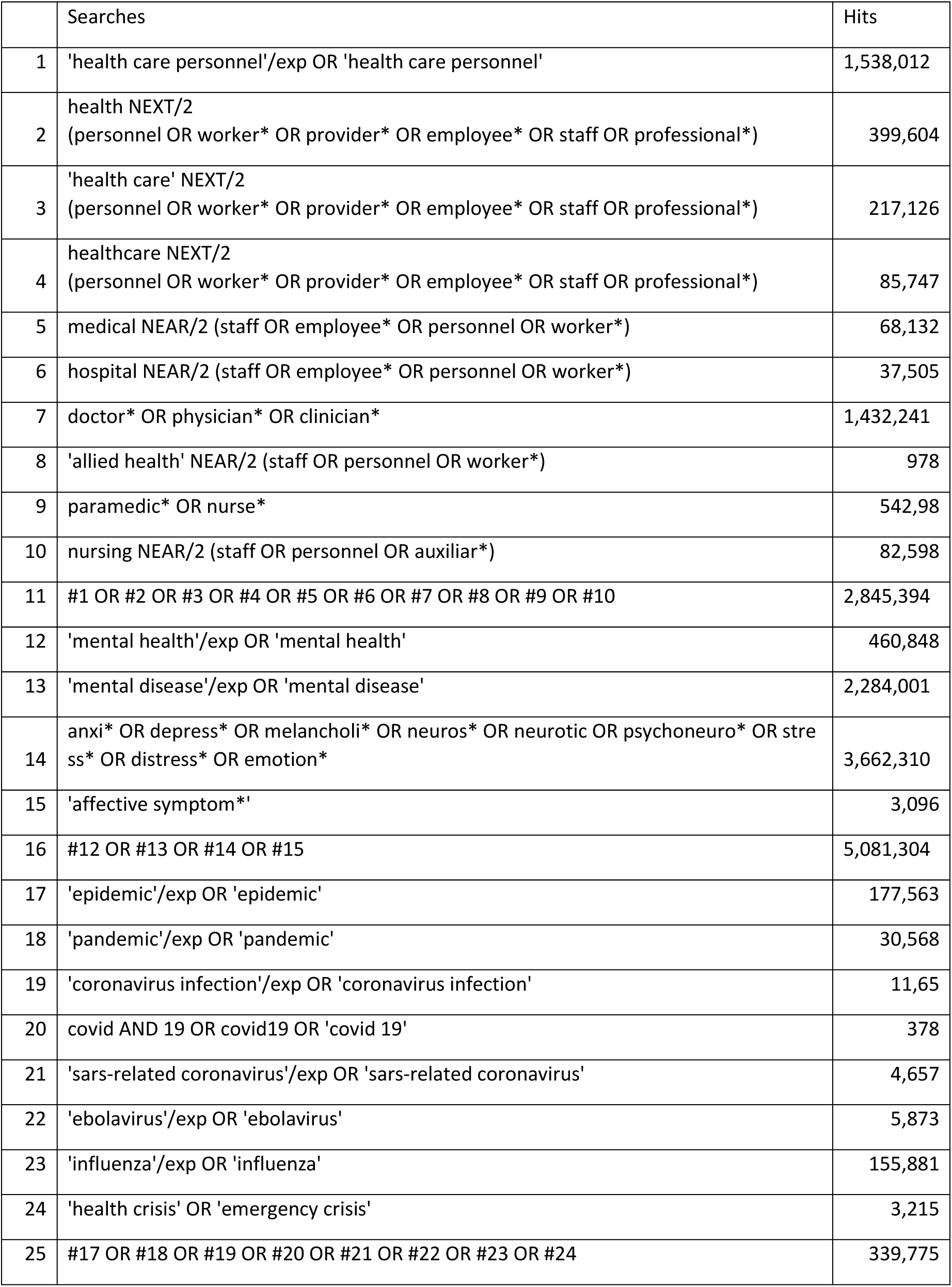
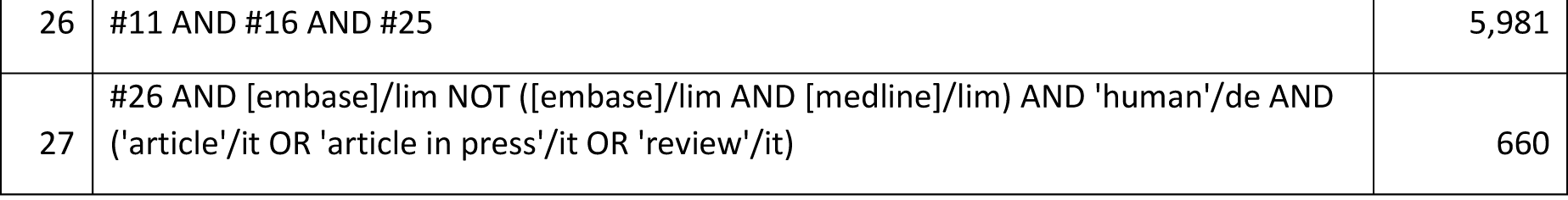

## Online Appendix 2 List of excluded papers after full-text screening

## Online Appendix 3 Risk of bias assessment

**A. Risk of bias of cross-sectional, assessed with the “Risk of Bias Instrument for Cross-Sectional Surveys of Attitudes and Practices” (Evidence Partners)**

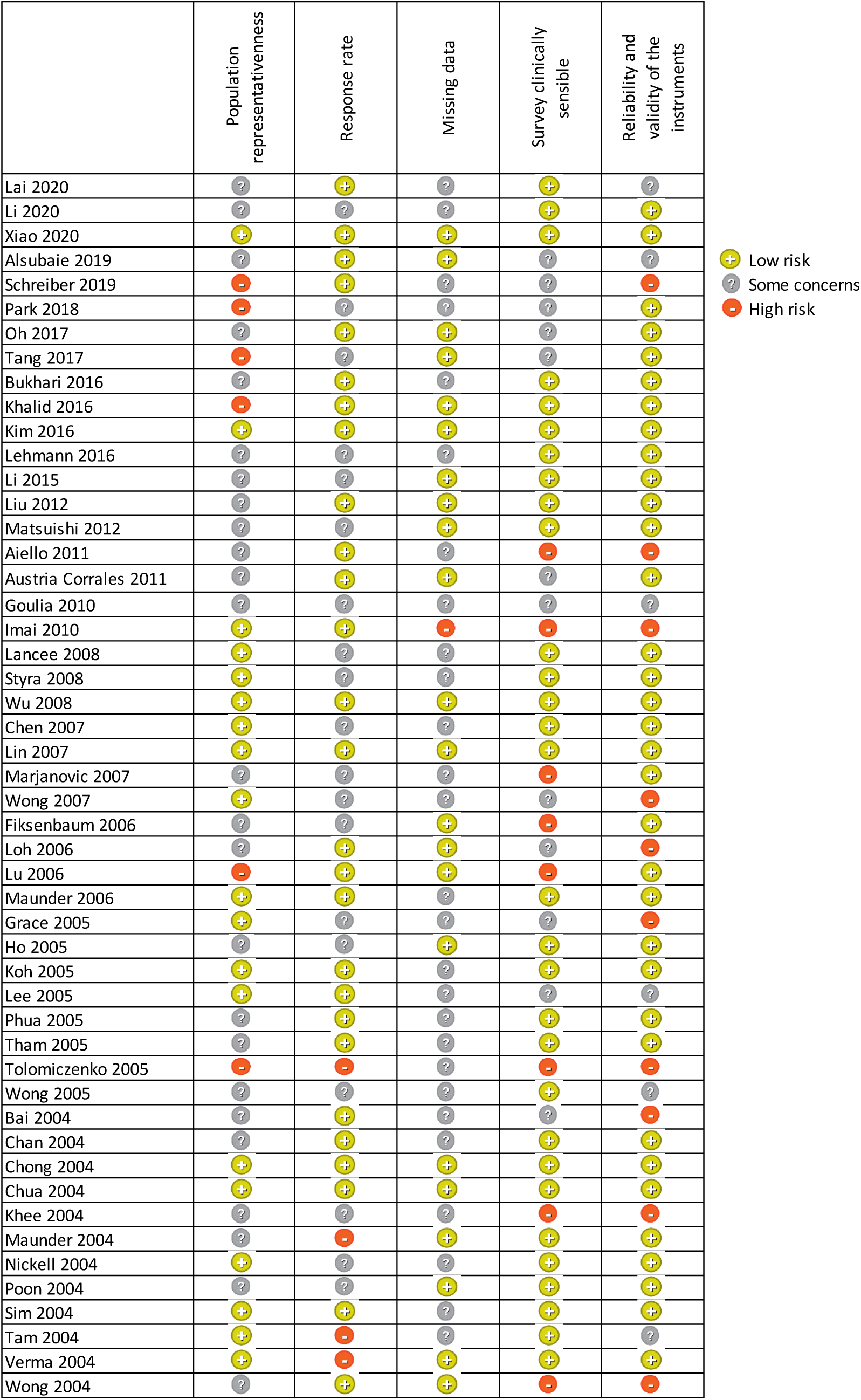

**B. Risk of bias of uncontrolled before-after studies (assess with ROBINS - I)**

|  | Bias due to confounding |  | Bias in selection of participants into the study |  | Bias in classification of interventions |  | Bias due to deviations from intended interventions |  | Bias due to missing data |  | Bias in measurement of outcomes |  | Bias in selection of the reported result |  | Overall bias |
| --- | --- | --- | --- | --- | --- | --- | --- | --- | --- | --- | --- | --- | --- | --- | --- |
|  | Rating | Comments | Rating | Comments | Rating | Comments | Rating | Comments | Rating | Comments | Rating | Comments | Rating | Comments |  |
| Chen 2006 | Low | GEE analysis for confounding factors and time-varying analysis. The negative side is the factors were self-reported. | Serious | Nurses volunteered to participate in the study (participation probably related to intervention/outcome). No preliminary analysis or assessment. | Moderate | (i) Intervention status is well defined; and (ii) Some aspects of the assignments of intervention status were determined retrospectively (participation was voluntary and knowledge of the prevention program may have affected the intervention). | No information | Not applicable. It is not possible to observe any differences from the intended intervention. | Low | - | Serious | Subjective measures. Team was aware of allocation status. | Moderate | GEE analysis is presented later in the results, not in the methods. | Serious |
| Maunderr 2010 | Critical | No adjusted analysis of confounding factors | Serious |  | Moderate | (i) Intervention status is well defined; and (ii) Some aspects of the assignments of intervention | No information | Not applicable. It is not possible to observe any differences from the intended | Serious | Proportions of missing participants differ substantially across interventions: Attrition was 12% in short | Serious | Subjective measures. Team was aware of allocation status. | Low | All outcomes listed in the methods are also reported in the results. Do not | Critical |
|  |  |  |  |  |  | status were determined retrospectively (participation was voluntary and knowledge of the prevention program may have affected the intervention) . |  | intervention. |  | courses and 28% in long courses.<br>The authors state a ITT in the methods, but no results were provided. |  |  |  | expect selective reporting . |  |

**C. Risk of bias of the systematic review identified (Brooks 2018), assessed with AMSTAR**.

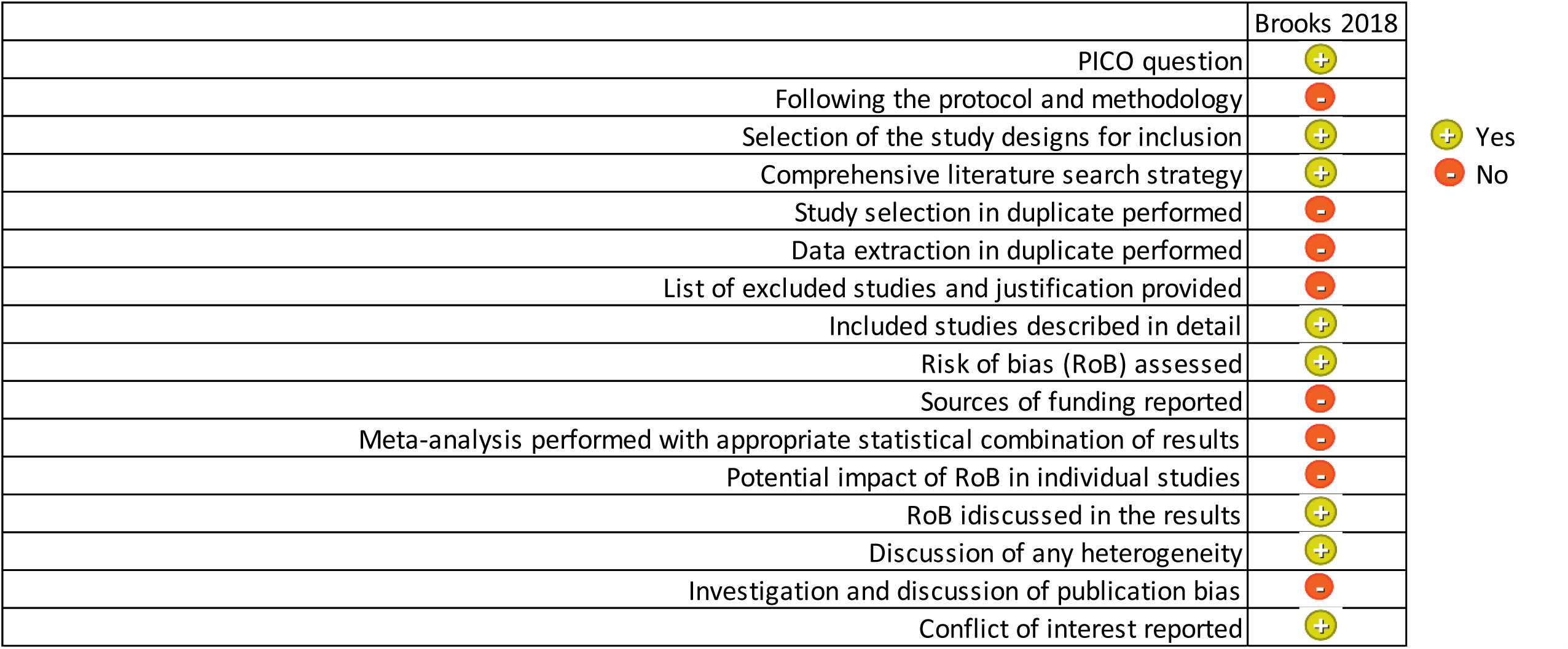

**D. Risk of bias of cohort studies, assessed with the “Tool to Assess Risk of Bias in Cohort Studies” (Evidence Partners)**

|  | Exposed and non-exposed cohorts drawn from the same population | Confidence in the assessment of exposure | Outcome not present at start of study | Associated or prognostic variables analyzed | Assessment of prognostic factors | Assessment of outcome | Follow up of cohorts | Co-interventions similar between groups |
| --- | --- | --- | --- | --- | --- | --- | --- | --- |
| Lee 2018 | - | + | ? | + | + | ? | ? | ? |
| Ji 2017 | - | + | ? | + | + | + | - | ? |
| Lung 2009 | + | + | - | + | + | + | ? | ? |
| Wu 2009 | ? | - | + | + | + | + | + | ? |
| Lee 2007 | + | + | ? | + | + | + | ? | ? |
| McAlonan 2007 | + | + | ? | + | ? | + | ? | ? |
| Su 2007 | + | + | - | + | + | + | + | + |

+ Low risk ? Some concerns - High risk

**E. Risk of bias of case control studies, assessed with the “Tool to Assess Risk of Bias in Case Control Studies” (Evidence Partners)**

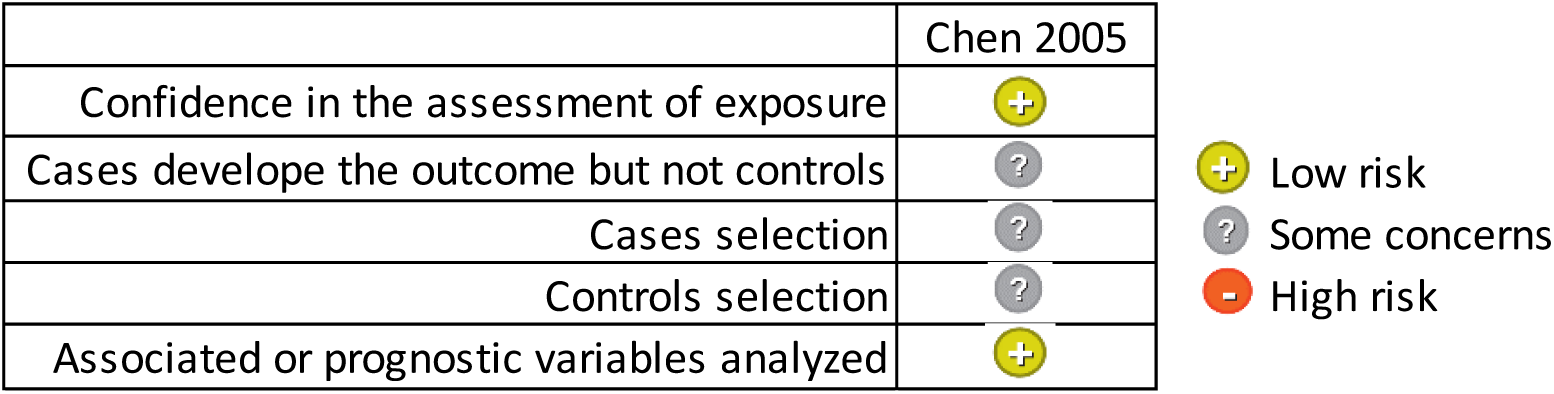

## Online Appendix 4 Summary of Findings table of studies examining the impact on mental health problems in healthcare workers during and after viral epidemics

**A. Summary of Findings table of studies examining the impact on mental health problems in healthcare workers during viral epidemics (N= 27)**

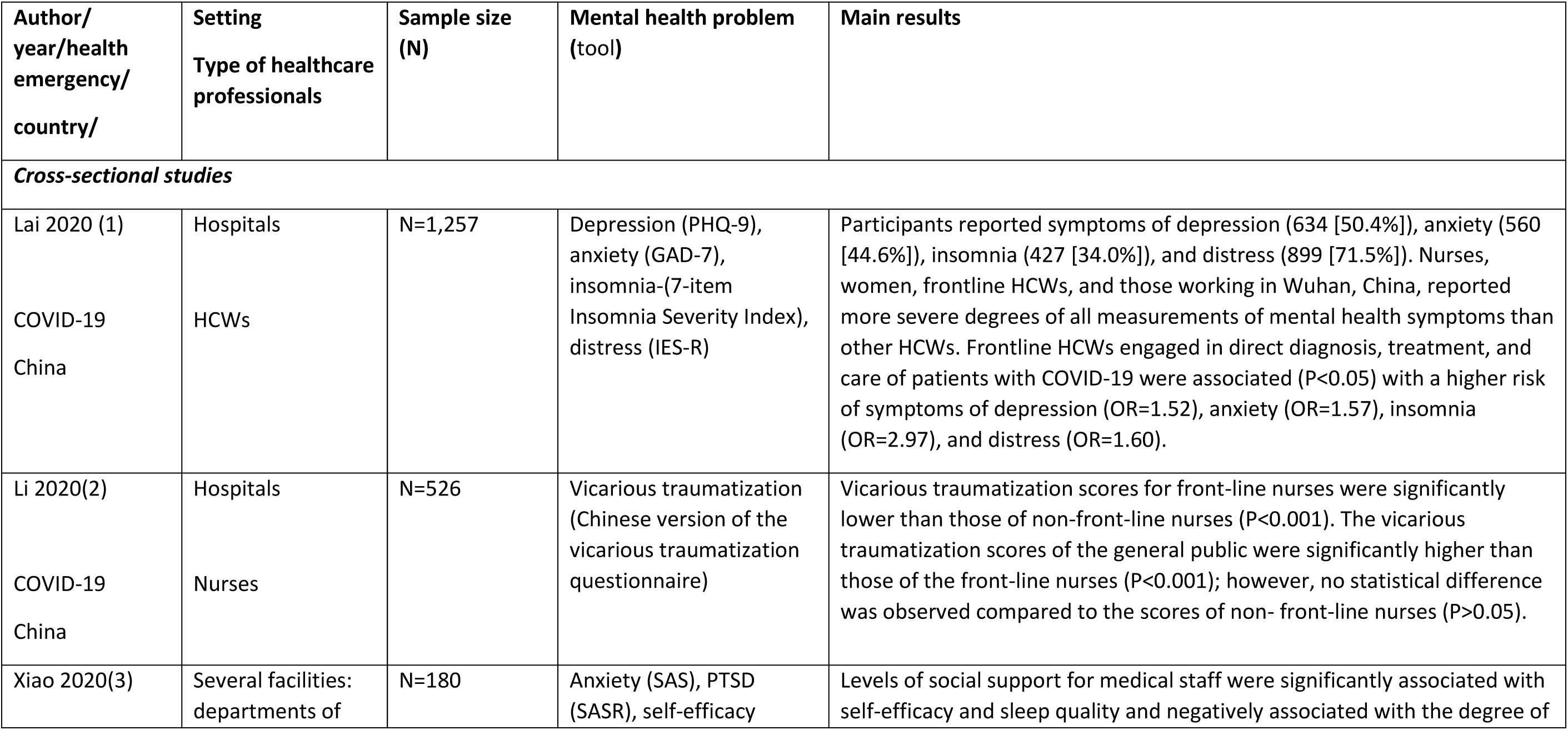
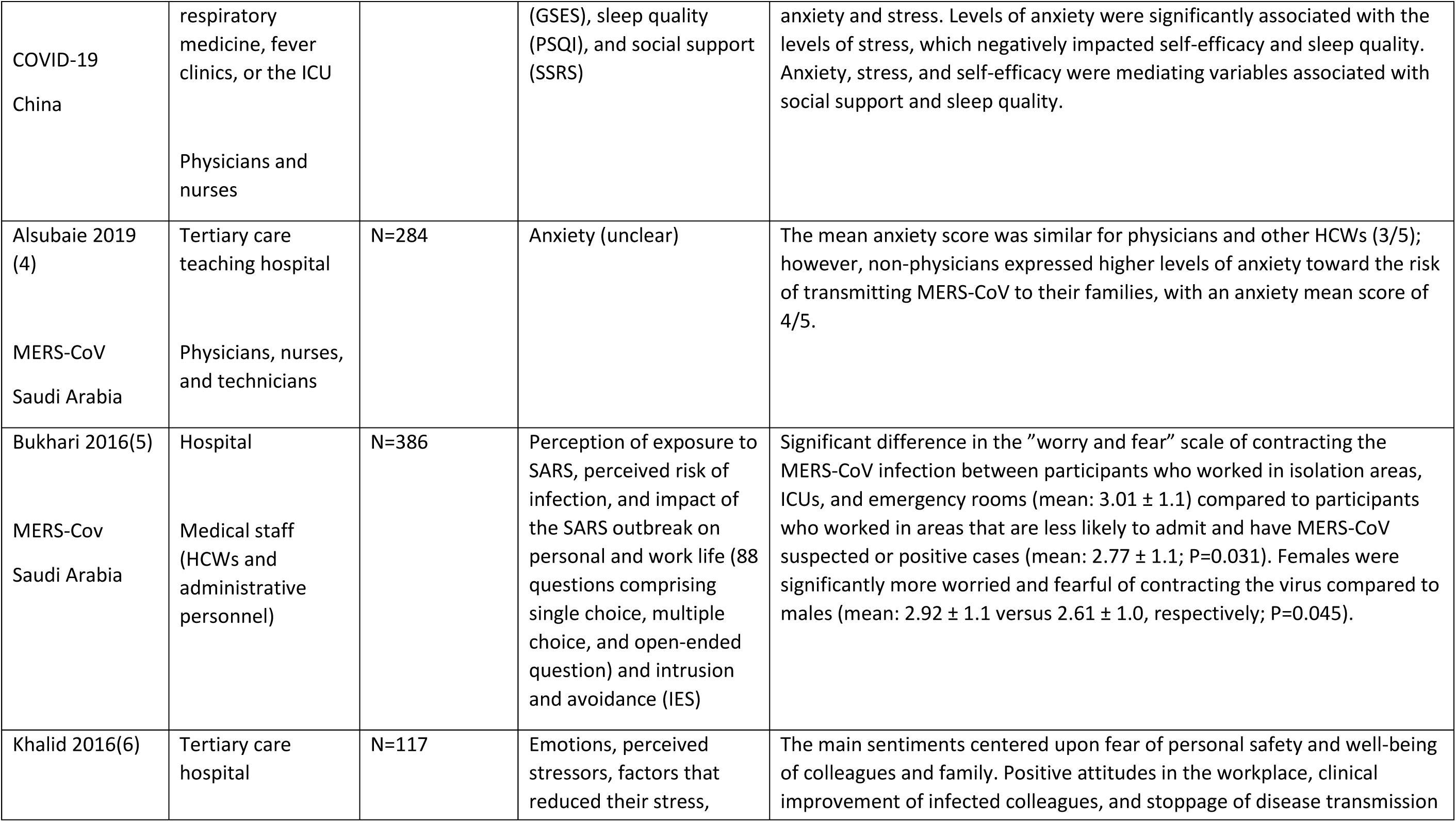
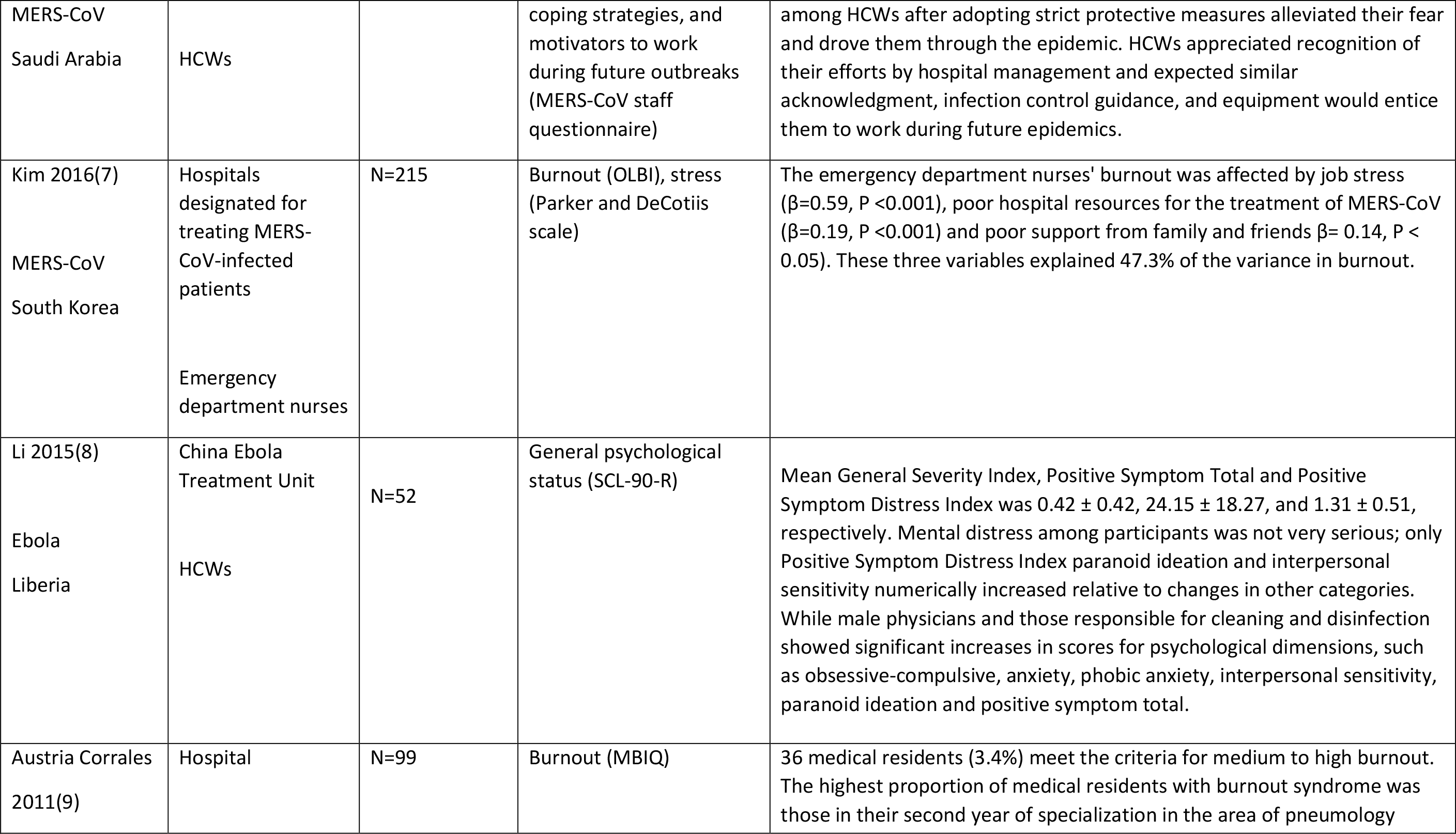
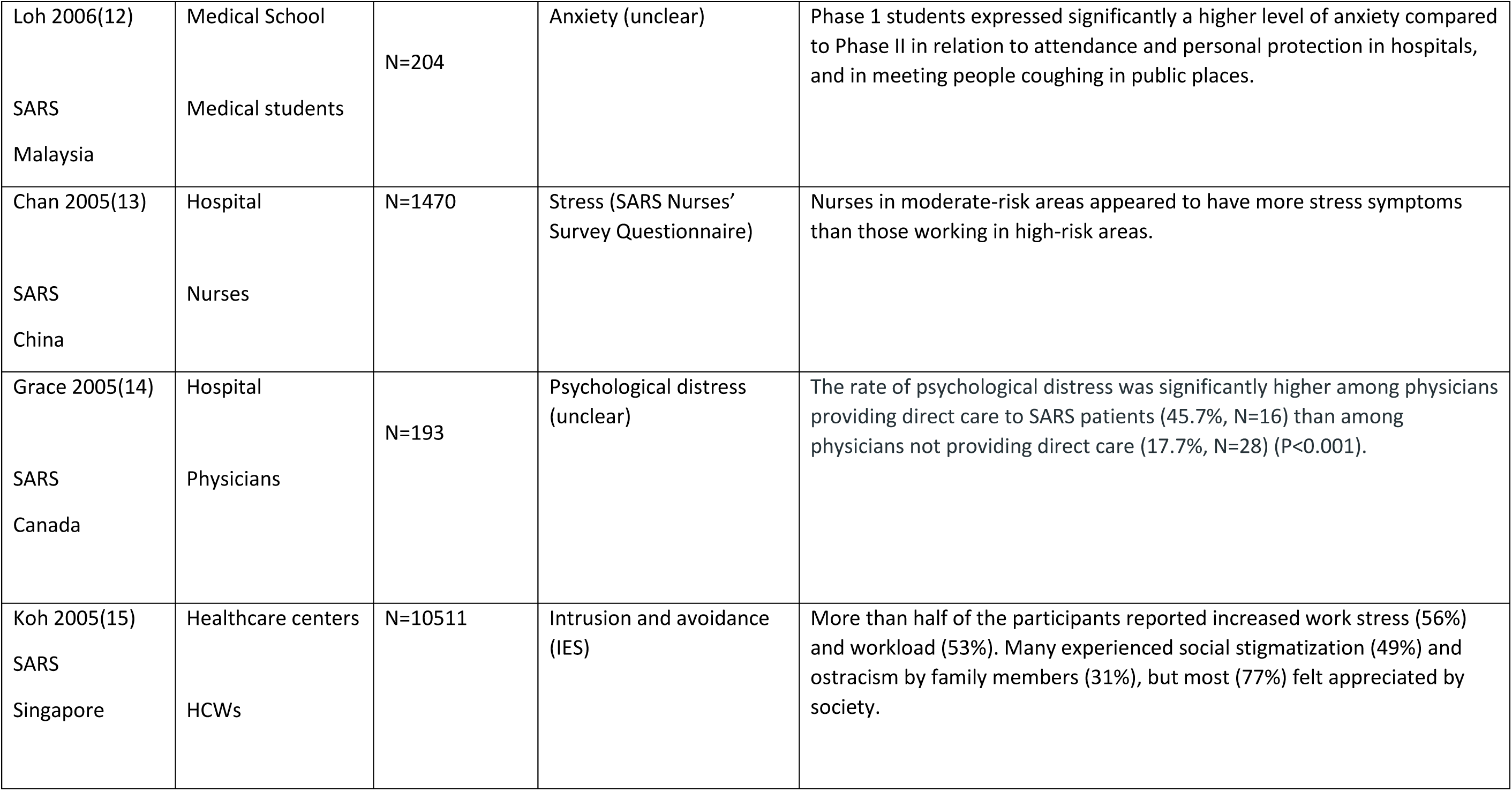
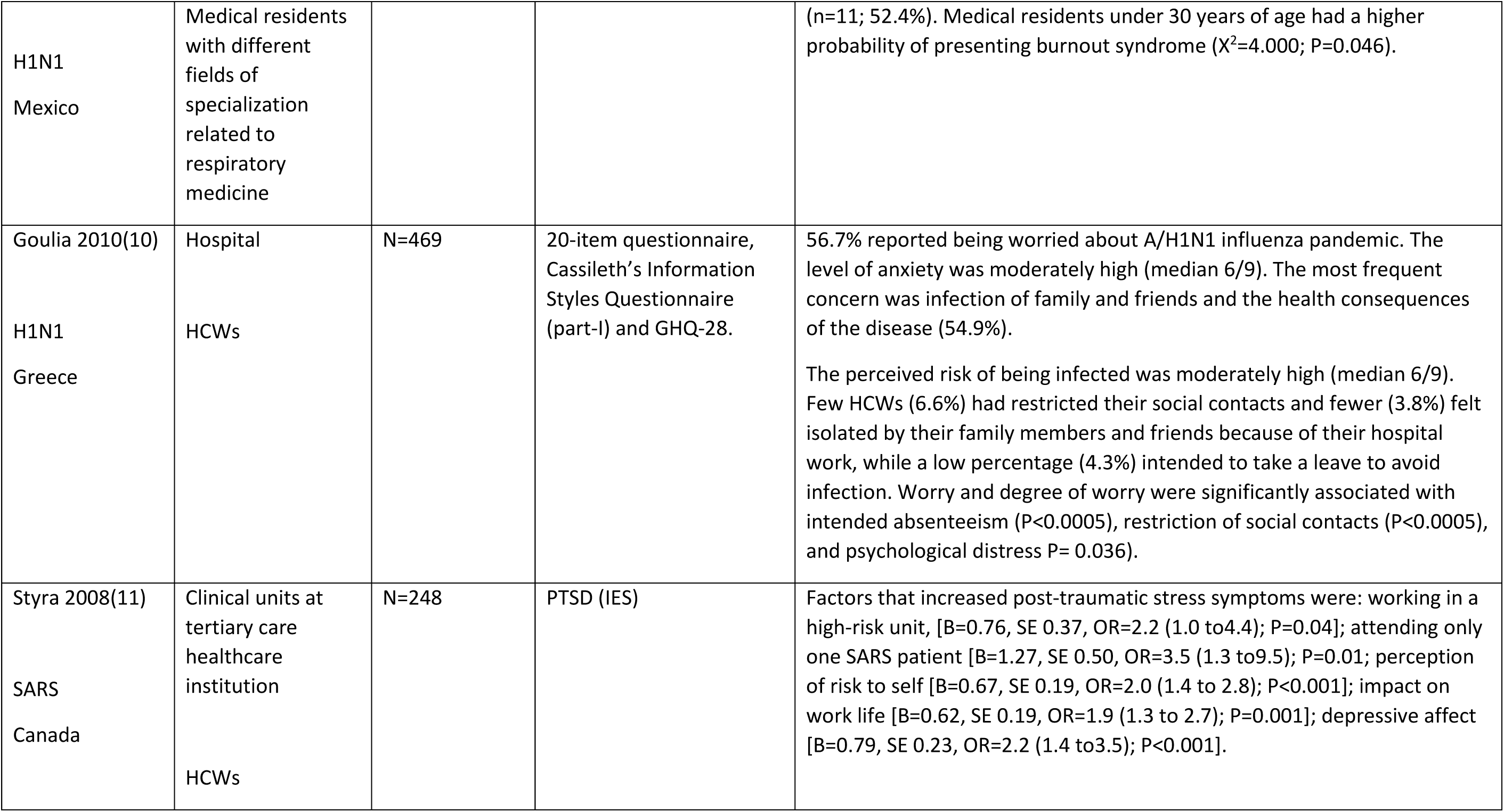
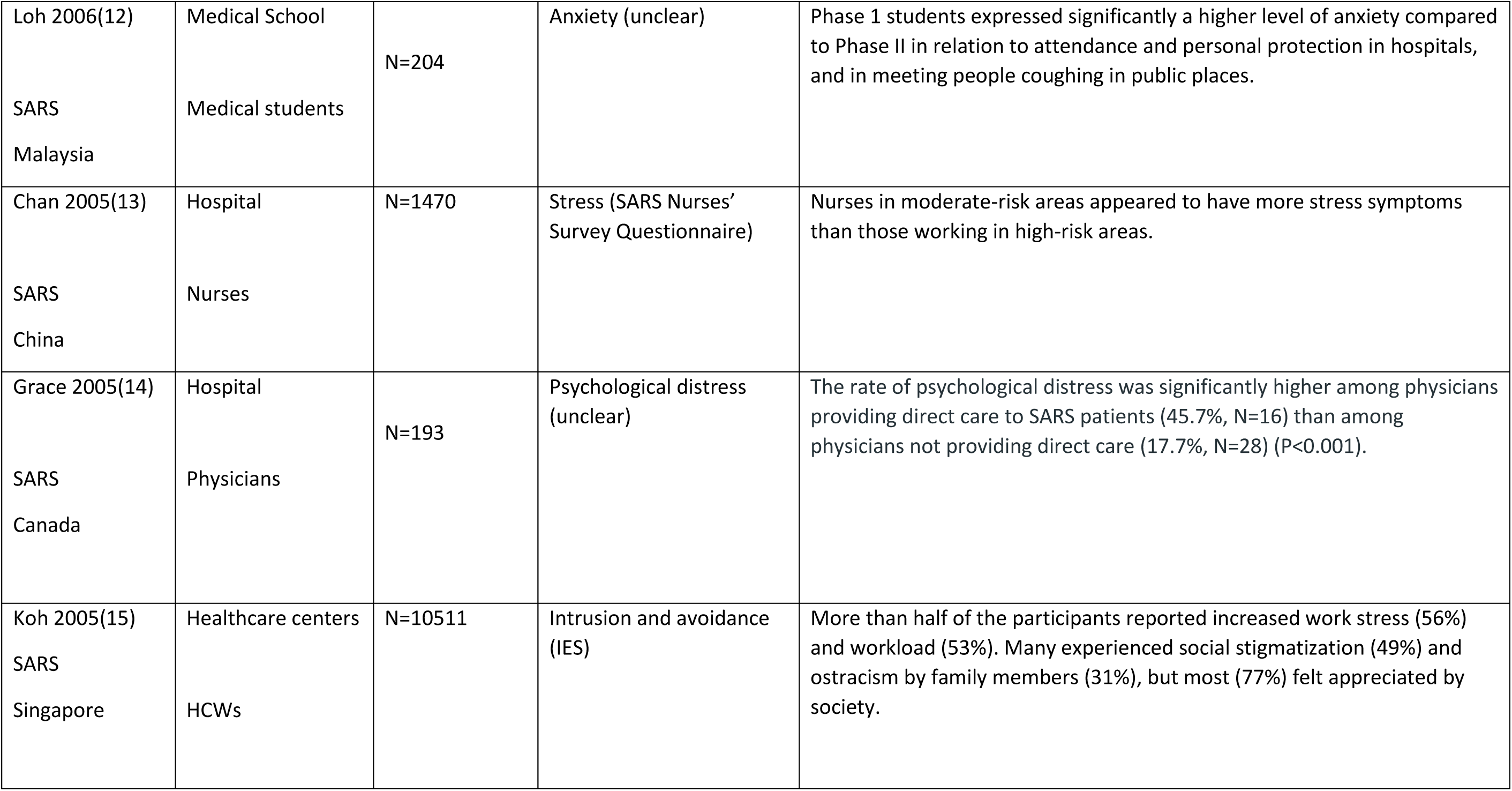
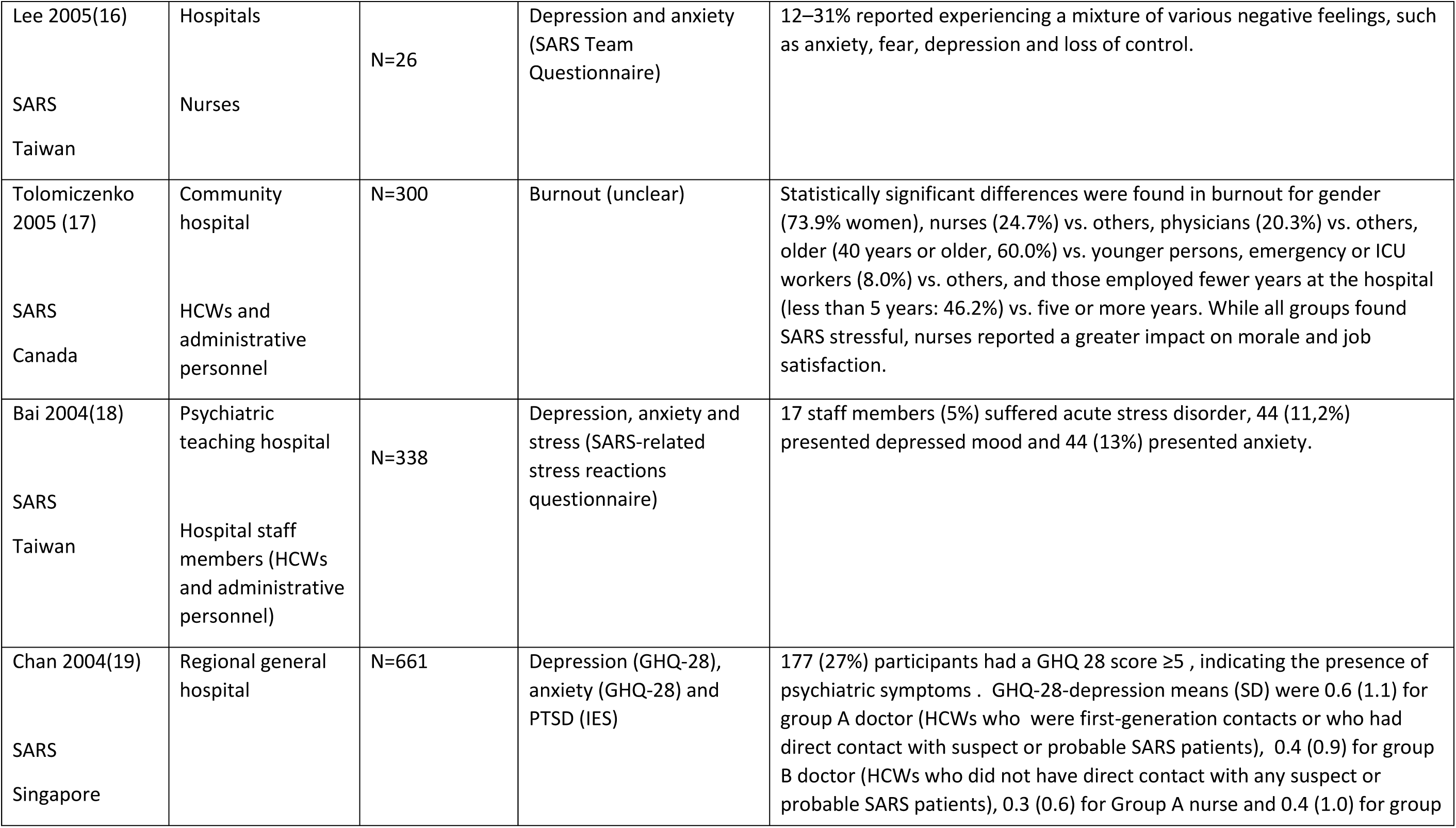
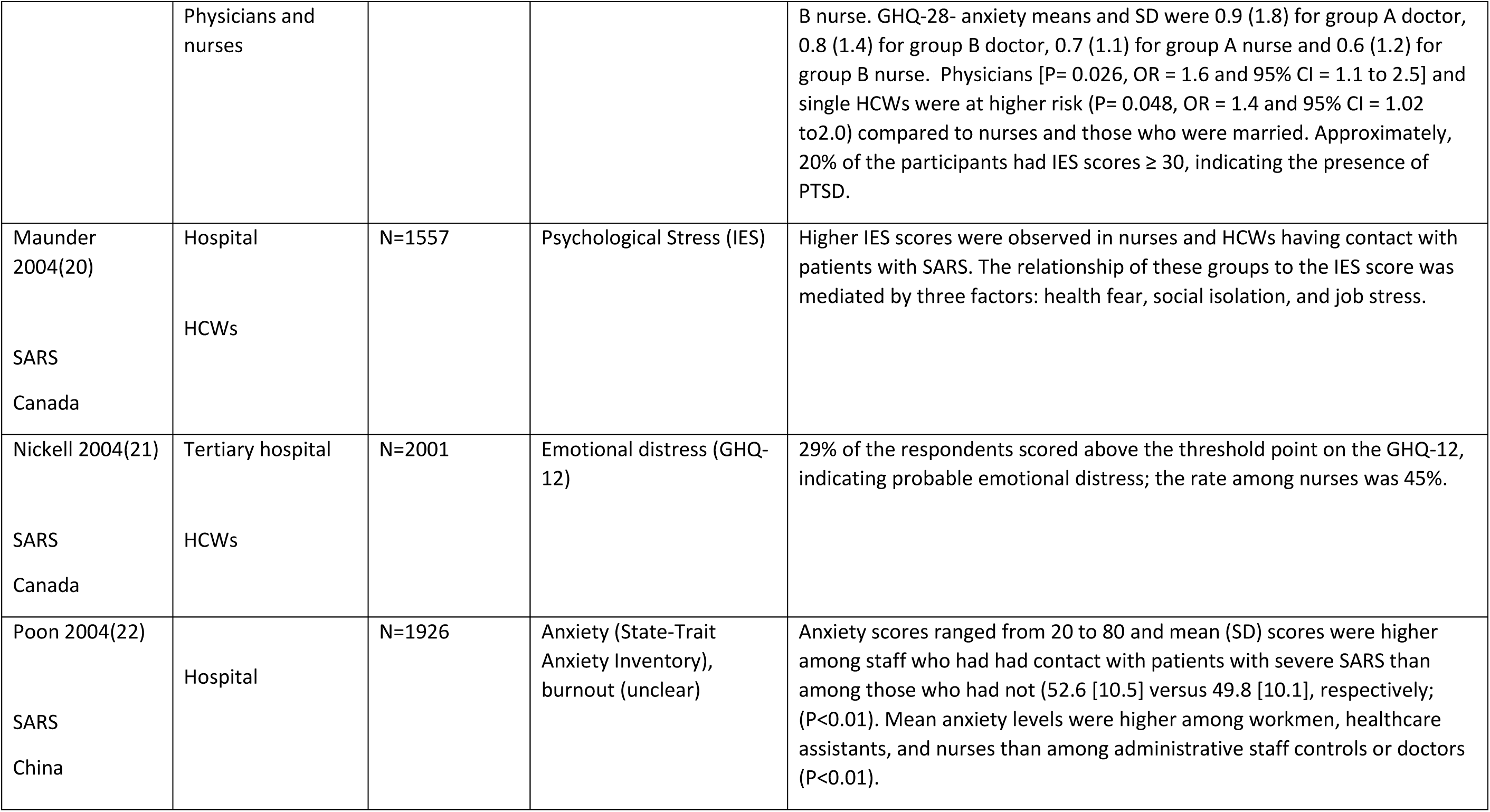
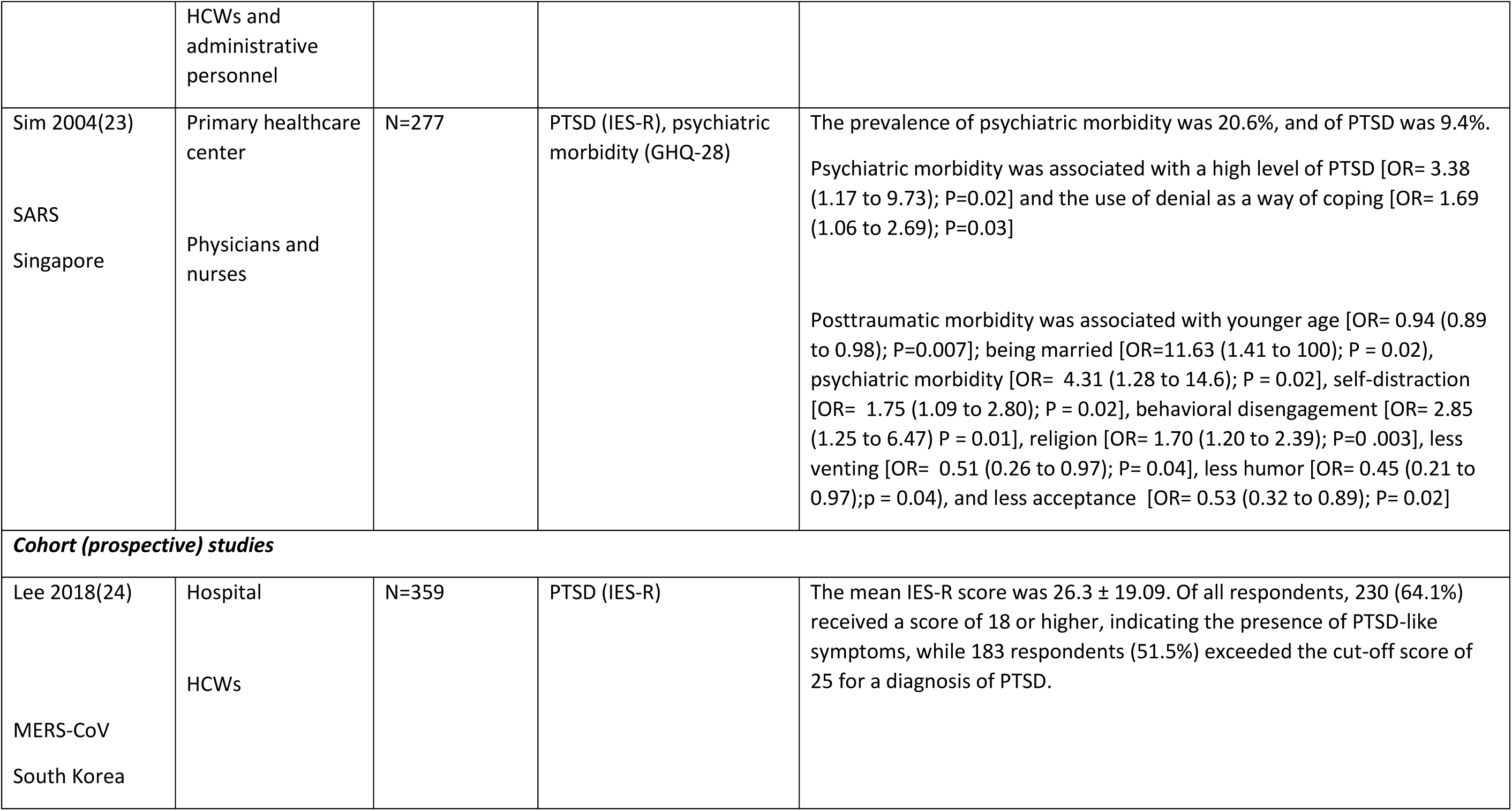
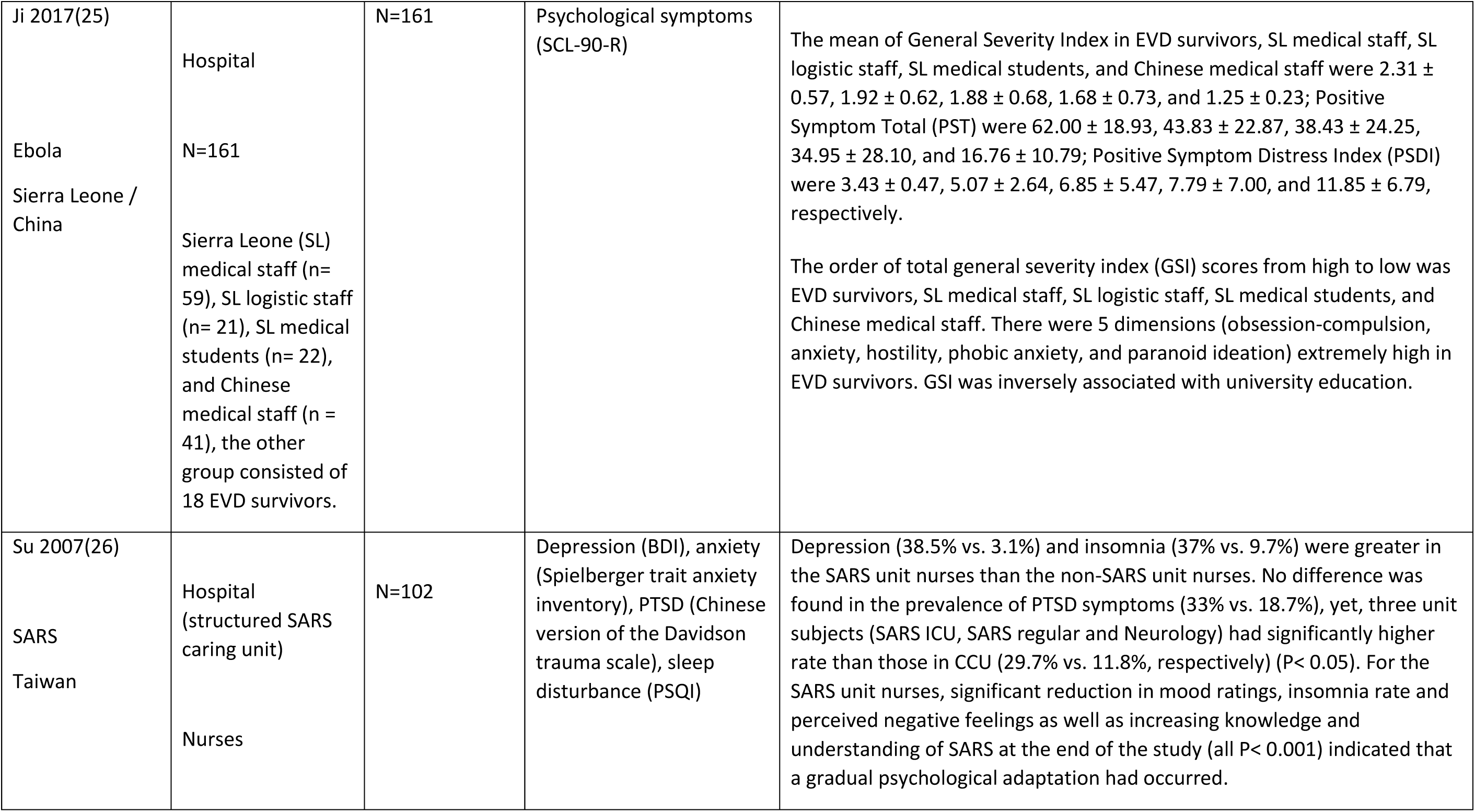
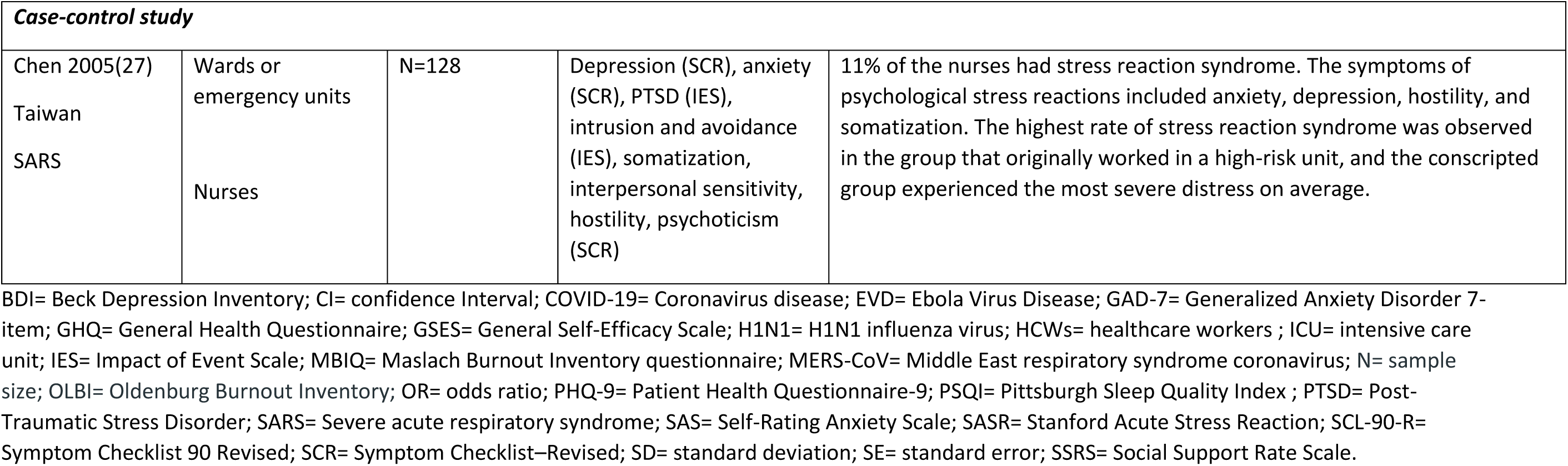

| **Author/**<br>**year/health**<br>**emergency/**<br>**country/** | **Setting**<br>**Type of healthcare**<br>**professionals** | **Sample size**<br>**(N)** | **Mental health problem**<br>**(tool)** | **Main results** |
| --- | --- | --- | --- | --- |
| ***Cross-sectional studies*** |  |  |  |  |
| Lai 2020 (1)<br>COVID-19<br>China | Hospitals<br>HCWs | N=1,257 | Depression (PHQ-9),<br>anxiety (GAD-7),<br>insomnia-(7-item<br>Insomnia Severity Index),<br>distress (IES-R) | Participants reported symptoms of depression (634 [50.4%]), anxiety (560 [44.6%]), insomnia (427 [34.0%]), and distress (899 [71.5%]). Nurses, women, frontline HCWs, and those working in Wuhan, China, reported more severe degrees of all measurements of mental health symptoms than other HCWs. Frontline HCWs engaged in direct diagnosis, treatment, and care of patients with COVID-19 were associated ($P<0.05$) with a higher risk of symptoms of depression ($OR=1.52$), anxiety ($OR=1.57$), insomnia ($OR=2.97$), and distress ($OR=1.60$). |
| Li 2020(2)<br>COVID-19<br>China | Hospitals<br>Nurses | N=526 | Vicarious traumatization<br>(Chinese version of the<br>vicarious traumatization<br>questionnaire) | Vicarious traumatization scores for front-line nurses were significantly lower than those of non-front-line nurses ($P<0.001$). The vicarious traumatization scores of the general public were significantly higher than those of the front-line nurses ($P<0.001$); however, no statistical difference was observed compared to the scores of non- front-line nurses ($P>0.05$). |
| Xiao 2020(3) | Several facilities:<br>departments of | N=180 | Anxiety (SAS), PTSD<br>(SASR), self-efficacy | Levels of social support for medical staff were significantly associated with self-efficacy and sleep quality and negatively associated with the degree of |
| COVID-19<br>China | respiratory medicine, fever clinics, or the ICU<br>Physicians and nurses |  | (GSES), sleep quality (PSQI), and social support (SSRS) | anxiety and stress. Levels of anxiety were significantly associated with the levels of stress, which negatively impacted self-efficacy and sleep quality. Anxiety, stress, and self-efficacy were mediating variables associated with social support and sleep quality. |
| Alsubaie 2019 (4)<br>MERS-CoV<br>Saudi Arabia | Tertiary care teaching hospital<br>Physicians, nurses, and technicians | N=284 | Anxiety (unclear) | The mean anxiety score was similar for physicians and other HCWs (3/5); however, non-physicians expressed higher levels of anxiety toward the risk of transmitting MERS-CoV to their families, with an anxiety mean score of 4/5. |
| Bukhari 2016(5)<br>MERS-Cov<br>Saudi Arabia | Hospital<br>Medical staff (HCWs and administrative personnel) | N=386 | Perception of exposure to SARS, perceived risk of infection, and impact of the SARS outbreak on personal and work life (88 questions comprising single choice, multiple choice, and open-ended question) and intrusion and avoidance (IES) | Significant difference in the "worry and fear" scale of contracting the MERS-CoV infection between participants who worked in isolation areas, ICUs, and emergency rooms (mean: $3.01 \pm 1.1$) compared to participants who worked in areas that are less likely to admit and have MERS-CoV suspected or positive cases (mean: $2.77 \pm 1.1$; $P=0.031$). Females were significantly more worried and fearful of contracting the virus compared to males (mean: $2.92 \pm 1.1$ versus $2.61 \pm 1.0$, respectively; $P=0.045$). |
| Khalid 2016(6) | Tertiary care hospital | N=117 | Emotions, perceived stressors, factors that reduced their stress, | The main sentiments centered upon fear of personal safety and well-being of colleagues and family. Positive attitudes in the workplace, clinical improvement of infected colleagues, and stoppage of disease transmission |
| MERS-CoV<br>Saudi Arabia | HCWs |  | coping strategies, and motivators to work during future outbreaks (MERS-CoV staff questionnaire) | among HCWs after adopting strict protective measures alleviated their fear and drove them through the epidemic. HCWs appreciated recognition of their efforts by hospital management and expected similar acknowledgment, infection control guidance, and equipment would entice them to work during future epidemics. |
| Kim 2016(7)<br>MERS-CoV<br>South Korea | Hospitals designated for treating MERS-CoV-infected patients<br>Emergency department nurses | N=215 | Burnout (OLBI), stress (Parker and DeCotiis scale) | The emergency department nurses' burnout was affected by job stress ($\beta=0.59$, $P < 0.001$), poor hospital resources for the treatment of MERS-CoV ($\beta=0.19$, $P < 0.001$) and poor support from family and friends $\beta= 0.14$, $P < 0.05$). These three variables explained 47.3% of the variance in burnout. |
| Li 2015(8)<br>Ebola<br>Liberia | China Ebola Treatment Unit<br>HCWs | N=52 | General psychological status (SCL-90-R) | Mean General Severity Index, Positive Symptom Total and Positive Symptom Distress Index was $0.42 \pm 0.42$, $24.15 \pm 18.27$, and $1.31 \pm 0.51$, respectively. Mental distress among participants was not very serious; only Positive Symptom Distress Index paranoid ideation and interpersonal sensitivity numerically increased relative to changes in other categories. While male physicians and those responsible for cleaning and disinfection showed significant increases in scores for psychological dimensions, such as obsessive-compulsive, anxiety, phobic anxiety, interpersonal sensitivity, paranoid ideation and positive symptom total. |
| Austria Corrales<br>2011(9) | Hospital | N=99 | Burnout (MBIQ) | 36 medical residents (3.4%) meet the criteria for medium to high burnout. The highest proportion of medical residents with burnout syndrome was those in their second year of specialization in the area of pneumology |
| H1N1<br>Mexico | Medical residents with different fields of specialization related to respiratory medicine | | | (n=11; 52.4%). Medical residents under 30 years of age had a higher probability of presenting burnout syndrome ($X^2=4.000$; $P=0.046$). |
| Goulia 2010(10)<br>H1N1<br>Greece | Hospital<br>HCWs | N=469 | 20-item questionnaire, Cassileth's Information Styles Questionnaire (part-I) and GHQ-28. | 56.7% reported being worried about A/H1N1 influenza pandemic. The level of anxiety was moderately high (median 6/9). The most frequent concern was infection of family and friends and the health consequences of the disease (54.9%).<br>The perceived risk of being infected was moderately high (median 6/9). Few HCWs (6.6%) had restricted their social contacts and fewer (3.8%) felt isolated by their family members and friends because of their hospital work, while a low percentage (4.3%) intended to take a leave to avoid infection. Worry and degree of worry were significantly associated with intended absenteeism ($P<0.0005$), restriction of social contacts ($P<0.0005$), and psychological distress $P=0.036$. |
| Styra 2008(11)<br>SARS<br>Canada | Clinical units at tertiary care healthcare institution<br>HCWs | N=248 | PTSD (IES) | Factors that increased post-traumatic stress symptoms were: working in a high-risk unit, $[B=0.76, SE 0.37, OR=2.2 (1.0 \text{ to } 4.4); P=0.04]$; attending only one SARS patient $[B=1.27, SE 0.50, OR=3.5 (1.3 \text{ to } 9.5); P=0.01]$; perception of risk to self $[B=0.67, SE 0.19, OR=2.0 (1.4 \text{ to } 2.8); P<0.001]$; impact on work life $[B=0.62, SE 0.19, OR=1.9 (1.3 \text{ to } 2.7); P=0.001]$; depressive affect $[B=0.79, SE 0.23, OR=2.2 (1.4 \text{ to } 3.5); P<0.001]$. |
| Loh 2006(12) | Medical School | N=204 | Anxiety (unclear) | Phase 1 students expressed significantly a higher level of anxiety compared to Phase II in relation to attendance and personal protection in hospitals, and in meeting people coughing in public places. |
| SARS<br>Malaysia | Medical students |  |  |  |
| Chan 2005(13) | Hospital | N=1470 | Stress (SARS Nurses' Survey Questionnaire) | Nurses in moderate-risk areas appeared to have more stress symptoms than those working in high-risk areas. |
| SARS<br>China | Nurses |  |  |  |
| Grace 2005(14) | Hospital | N=193 | Psychological distress (unclear) | The rate of psychological distress was significantly higher among physicians providing direct care to SARS patients (45.7%, N=16) than among physicians not providing direct care (17.7%, N=28) ($P<0.001$). |
| SARS<br>Canada | Physicians |  |  |  |
| Koh 2005(15) | Healthcare centers | N=10511 | Intrusion and avoidance (IES) | More than half of the participants reported increased work stress (56%) and workload (53%). Many experienced social stigmatization (49%) and ostracism by family members (31%), but most (77%) felt appreciated by society. |
| SARS<br>Singapore | HCWs |  |  |  |
| Lee 2005(16)<br>SARS<br>Taiwan | Hospitals<br>Nurses | N=26 | Depression and anxiety (SARS Team Questionnaire) | 12–31% reported experiencing a mixture of various negative feelings, such as anxiety, fear, depression and loss of control. |
| Tolomiczenko 2005 (17)<br>SARS<br>Canada | Community hospital<br>HCWs and administrative personnel | N=300 | Burnout (unclear) | Statistically significant differences were found in burnout for gender (73.9% women), nurses (24.7%) vs. others, physicians (20.3%) vs. others, older (40 years or older, 60.0%) vs. younger persons, emergency or ICU workers (8.0%) vs. others, and those employed fewer years at the hospital (less than 5 years: 46.2%) vs. five or more years. While all groups found SARS stressful, nurses reported a greater impact on morale and job satisfaction. |
| Bai 2004(18)<br>SARS<br>Taiwan | Psychiatric teaching hospital<br>Hospital staff members (HCWs and administrative personnel) | N=338 | Depression, anxiety and stress (SARS-related stress reactions questionnaire) | 17 staff members (5%) suffered acute stress disorder, 44 (11,2%) presented depressed mood and 44 (13%) presented anxiety. |
| Chan 2004(19)<br>SARS<br>Singapore | Regional general hospital | N=661 | Depression (GHQ-28), anxiety (GHQ-28) and PTSD (IES) | 177 (27%) participants had a GHQ 28 score $\geq 5$ , indicating the presence of psychiatric symptoms . GHQ-28-depression means (SD) were 0.6 (1.1) for group A doctor (HCWs who were first-generation contacts or who had direct contact with suspect or probable SARS patients), 0.4 (0.9) for group B doctor (HCWs who did not have direct contact with any suspect or probable SARS patients), 0.3 (0.6) for Group A nurse and 0.4 (1.0) for group |
| | Physicians and nurses | | | B nurse. GHQ-28- anxiety means and SD were 0.9 (1.8) for group A doctor, 0.8 (1.4) for group B doctor, 0.7 (1.1) for group A nurse and 0.6 (1.2) for group B nurse. Physicians [P= 0.026, OR = 1.6 and 95% CI = 1.1 to 2.5] and single HCWs were at higher risk (P= 0.048, OR = 1.4 and 95% CI = 1.02 to 2.0) compared to nurses and those who were married. Approximately, 20% of the participants had IES scores $\geq 30$, indicating the presence of PTSD. |
| Maunder 2004(20)<br>SARS<br>Canada | Hospital<br>HCWs | N=1557 | Psychological Stress (IES) | Higher IES scores were observed in nurses and HCWs having contact with patients with SARS. The relationship of these groups to the IES score was mediated by three factors: health fear, social isolation, and job stress. |
| Nickell 2004(21)<br>SARS<br>Canada | Tertiary hospital<br>HCWs | N=2001 | Emotional distress (GHQ-12) | 29% of the respondents scored above the threshold point on the GHQ-12, indicating probable emotional distress; the rate among nurses was 45%. |
| Poon 2004(22)<br>SARS<br>China | Hospital | N=1926 | Anxiety (State-Trait Anxiety Inventory), burnout (unclear) | Anxiety scores ranged from 20 to 80 and mean (SD) scores were higher among staff who had had contact with patients with severe SARS than among those who had not (52.6 [10.5] versus 49.8 [10.1], respectively; (P<0.01). Mean anxiety levels were higher among workmen, healthcare assistants, and nurses than among administrative staff controls or doctors (P<0.01). |
|  | HCWs and administrative personnel |  |  |  |
| Sim 2004(23)<br>SARS<br>Singapore | Primary healthcare center<br>Physicians and nurses | N=277 | PTSD (IES-R), psychiatric morbidity (GHQ-28) | The prevalence of psychiatric morbidity was 20.6%, and of PTSD was 9.4%.<br>Psychiatric morbidity was associated with a high level of PTSD [OR= 3.38 (1.17 to 9.73); P=0.02] and the use of denial as a way of coping [OR= 1.69 (1.06 to 2.69); P=0.03]<br>Posttraumatic morbidity was associated with younger age [OR= 0.94 (0.89 to 0.98); P=0.007]; being married [OR=11.63 (1.41 to 100); P = 0.02], psychiatric morbidity [OR= 4.31 (1.28 to 14.6); P = 0.02], self-distraction [OR= 1.75 (1.09 to 2.80); P = 0.02], behavioral disengagement [OR= 2.85 (1.25 to 6.47) P = 0.01], religion [OR= 1.70 (1.20 to 2.39); P=0 .003], less venting [OR= 0.51 (0.26 to 0.97); P= 0.04], less humor [OR= 0.45 (0.21 to 0.97);p = 0.04], and less acceptance [OR= 0.53 (0.32 to 0.89); P= 0.02] |
| ***Cohort (prospective) studies*** |  |  |  |  |
| Lee 2018(24)<br>MERS-CoV<br>South Korea | Hospital<br>HCWs | N=359 | PTSD (IES-R) | The mean IES-R score was 26.3 ± 19.09. Of all respondents, 230 (64.1%) received a score of 18 or higher, indicating the presence of PTSD-like symptoms, while 183 respondents (51.5%) exceeded the cut-off score of 25 for a diagnosis of PTSD. |
| Ji 2017(25)<br>Ebola<br>Sierra Leone / China | Hospital<br>N=161<br>Sierra Leone (SL) medical staff (n= 59), SL logistic staff (n= 21), SL medical students (n= 22), and Chinese medical staff (n = 41), the other group consisted of 18 EVD survivors. | N=161 | Psychological symptoms (SCL-90-R) | The mean of General Severity Index in EVD survivors, SL medical staff, SL logistic staff, SL medical students, and Chinese medical staff were $2.31 \pm 0.57$, $1.92 \pm 0.62$, $1.88 \pm 0.68$, $1.68 \pm 0.73$, and $1.25 \pm 0.23$; Positive Symptom Total (PST) were $62.00 \pm 18.93$, $43.83 \pm 22.87$, $38.43 \pm 24.25$, $34.95 \pm 28.10$, and $16.76 \pm 10.79$; Positive Symptom Distress Index (PSDI) were $3.43 \pm 0.47$, $5.07 \pm 2.64$, $6.85 \pm 5.47$, $7.79 \pm 7.00$, and $11.85 \pm 6.79$, respectively.<br>The order of total general severity index (GSI) scores from high to low was EVD survivors, SL medical staff, SL logistic staff, SL medical students, and Chinese medical staff. There were 5 dimensions (obsession-compulsion, anxiety, hostility, phobic anxiety, and paranoid ideation) extremely high in EVD survivors. GSI was inversely associated with university education. |
| Su 2007(26)<br>SARS<br>Taiwan | Hospital (structured SARS caring unit)<br>Nurses | N=102 | Depression (BDI), anxiety (Spielberger trait anxiety inventory), PTSD (Chinese version of the Davidson trauma scale), sleep disturbance (PSQI) | Depression (38.5% vs. 3.1%) and insomnia (37% vs. 9.7%) were greater in the SARS unit nurses than the non-SARS unit nurses. No difference was found in the prevalence of PTSD symptoms (33% vs. 18.7%), yet, three unit subjects (SARS ICU, SARS regular and Neurology) had significantly higher rate than those in CCU (29.7% vs. 11.8%, respectively) ($P < 0.05$). For the SARS unit nurses, significant reduction in mood ratings, insomnia rate and perceived negative feelings as well as increasing knowledge and understanding of SARS at the end of the study (all $P < 0.001$) indicated that a gradual psychological adaptation had occurred. |

| **Case-control study** |  |  |  |  |
| --- | --- | --- | --- | --- |
| Chen 2005(27)<br>Taiwan<br>SARS | Wards or<br>emergency units<br>Nurses | N=128 | Depression (SCR), anxiety (SCR), PTSD (IES), intrusion and avoidance (IES), somatization, interpersonal sensitivity, hostility, psychoticism (SCR) | 11% of the nurses had stress reaction syndrome. The symptoms of psychological stress reactions included anxiety, depression, hostility, and somatization. The highest rate of stress reaction syndrome was observed in the group that originally worked in a high-risk unit, and the conscripted group experienced the most severe distress on average. |

BDI= Beck Depression Inventory; CI= confidence Interval; COVID-19= Coronavirus disease; EVD= Ebola Virus Disease; GAD-7= Generalized Anxiety Disorder 7-item; GHQ= General Health Questionnaire; GSES= General Self-Efficacy Scale; H1N1= H1N1 influenza virus; HCWs= healthcare workers ; ICU= intensive care unit; IES= Impact of Event Scale; MBIQ= Maslach Burnout Inventory questionnaire; MERS-CoV= Middle East respiratory syndrome coronavirus; N= sample size; OLB= Oldenburg Burnout Inventory; OR= odds ratio; PHQ-9= Patient Health Questionnaire-9; PSQI= Pittsburgh Sleep Quality Index ; PTSD= Post-Traumatic Stress Disorder; SARS= Severe acute respiratory syndrome; SAS= Self-Rating Anxiety Scale; SASR= Stanford Acute Stress Reaction; SCL-90-R= Symptom Checklist 90 Revised; SCR= Symptom Checklist–Revised; SD= standard deviation; SE= standard error; SSRS= Social Support Rate Scale.

**B. Summary of Findings table of studies examining the impact on mental health problems in healthcare workers after viral epidemics (N= 27)**

| **Author/**<br>**year/health**<br>**emergency/**<br>**country/** | **Setting**<br>**Type of**<br>**healthcare**<br>**professionals** | **Sample size (N)** | **Mental health problem**<br>**(tool)** | **Main results** |
| --- | --- | --- | --- | --- |
| ***Cross-sectional studies*** |  |  |  |  |
| Park 2018(28)<br>MERS-Cov<br>South Korea | Hospital<br>Nurses | N=187 | Mental Health (SF-36);<br>Stress (PSS-10); Hardiness<br>(DSR-15); Stigma (new<br>scale for study) | Mental Health: a significant direct effect of stigma on mental health when<br>controlling for hardiness and stress ($\beta = -0.306$, $t = -7.2376$, $P < 0.001$)<br>Hardiness exerted a significant direct effect on mental health when the<br>other variables were kept constant ($\beta=0.487$, $t=4.8692$, $P<0.001$)<br>The indirect effect of stigma on mental health was 19.9% of its direct effect<br>on mental health, while the indirect effect of hardiness was 51.6% of its<br>direct effect |
| Oh 2017(29)<br>MERS-Cov<br>South Korea | Hospital<br>Nurses | N=313 | Stress (12 questions<br>based on a trauma<br>appraisal questionnaire<br>and questionnaire<br>developed by the Korean<br>Neuro-Psychiatric<br>Association for medical | The overall mean stress score of total study participants was 32.91.<br>Nurses who provided either inpatient nursing care or screening services of<br>suspected or confirmed case (first hand): overall stress= 33.72 |
|  |  |  | workers who experienced the MERS outbreak) | Nurses who participated in medical care of the general population with no suspected MERS symptom (second hand): overall stress= 32.25<br>(There was no statistically significant difference between the groups)<br>The regression estimate between stress and nursing intention was $B (SE)= -0.08 (0.02)$ $\beta= -0.21$ $P<0.01$. |
| Tang 2017(30) | Departments admitting H7N9 patients | N=102 | PTSD (PCL-C) | 20.59% showed PTSD symptoms. The sample had a mean PCL-C score of $30\pm9.95$. The differences in the scores of doctors and nurses with different genders, ages, professional titles, contact frequencies, trainings, and experiences were statistically significant ($P<0.05$, $P<0.01$). Moreover, t-tests and one-way analysis of variance showed that nurses received higher scores than doctors, female participants received higher scores than male participants, and the participants with low professional title and high contact frequency, aged between 20 years and 30 years, with less than five years of work experience, having not received related training and with no related experience obtained higher PCL-C scores than the others ($P<0.05$, $P<0.01$). |
| H7N9 influenza<br>China | HCWs |  |  |  |
| Lehmann 2016(31) | Hospital and research institute | N=86 | Depression (PHQ-9), anxiety (GAD-7), general mental health (SF12) | Ebola patient treatment group experienced significantly higher levels of social isolation than both other groups. The best predictors of poor physical and mental HrQoL were perceived lack of knowledge about the Ebola virus disease (physical: $\beta=-1.2$, $P=0.05$; mental: $\beta=-1.3$, $P=0.03$) and fatigue (physical: $\beta=-0.3$, $P=0.02$; mental: $\beta=-0.53$, $P<0.001$). |
| Ebola<br>Germany | HCWs, and general staff |  |  |  |
| Liu 2012(32)<br>SARS<br>China | Hospital<br>HCWs | N=549 | Depression (CES-D - Chinese version), PTSD (IES-R - Adapted and validated in Chinese) | The results of multinomial regression analyses showed that, with other relevant factors controlled for, being single, having been quarantined during the outbreak, having been exposed to other traumatic events before SARS, and perceived SARS-related risk level during the outbreak were found to increase the odds of having a high level of depressive symptoms 3 years later. Altruistic acceptance of risk during the outbreak was found to decrease the odds of high post-outbreak depressive symptom levels |
| Matsuishi 2012(33)<br>H1N1<br>Japan | Hospital<br>HCWs | N=1625 | Anxiety (19 stress-related questions), PTSD (IES) | Workers at a hospital with intense liaison psychiatric services felt less psychological impact. Workers at a hospital that provided staff with information about the pandemic less frequently, felt unprotected. Workers in work environments that had a high risk of infection felt more anxious and more exhausted. The total IES score was higher in workers in high-risk work environments. |
| Lancee 2008(34)<br>SARS<br>Canada | Academic and community hospitals<br>HCWs | N=139 | Depression, anxiety (Kessler Psychological Distress Scale), PTSD (IES), burnout (MBI) | The lifetime prevalence of any depressive, anxiety, or substance use diagnosis was 30%. Only one HCW who identified the SARS experience as a traumatic event was diagnosed as having PTSD. New episodes of psychiatric disorders occurred among seven healthcare workers (5%). New episodes of psychiatric disorders were directly associated with a history of having a psychiatric disorder before the SARS outbreak ($P=0.02$) and inversely associated with years of healthcare experience ($P=0.03$) and the perceived adequacy of training and support ($P=0.03$). |
| Wu 2008(35)<br>SARS<br>China | Hospital<br>HCWs | N=549 | PTSD (IES-R) | Current alcohol abuse/dependence symptom counts 3 years after the outbreak were positively associated with having been quarantined, or worked in high-risk locations such as SARS wards, during the outbreak. However, having had family members or friends contract, SARS was not related to alcohol abuse/dependence symptom count. Symptoms of PTS and of depression, and having used drinking as a coping method, were also significantly associated with increased alcohol abuse/dependence symptoms. The relationship between outbreak exposure and alcohol abuse/dependence symptom count remained significant even when sociodemographic and other factors were controlled for. When the intrusion, avoidance and hyperarousal PTS symptom clusters were entered into the model, hyperarousal was found to be significantly associated with alcohol abuse/dependence symptoms |
| Chen 2007 (36)<br>SARS<br>Taiwan | Tertiary care referral center<br>66 (73.3%) critical care nurses, 11 (12.2%) physicians, 7 (7.8%) technicians, and 6 (6.7%) respiratory care specialists. | N=172 | Role emotional, and mental health: MOS SF-36 | All serum specimens tested negative for SARS antibody. Survey scores for SARS HCWs immediately after care were significantly lower than those for the control group ($P < 0.05$ by the t test) in 6 categories. Vitality, social functioning, and mental health immediately after care and vitality and mental health after self-quarantine and off-duty shifts were among the worst subscales. The social functioning, role emotional, and role physical subscales significantly improved after self-quarantine and off-duty shifts ($P < 0.05$, by paired t test). The length of contact time (mean number of contact-hours per day) with patients with SARS was associated with some subscales (role emotional, role physical, and mental health) to a mild extent. The total number of contact-hours with symptomatic patients with SARS was a borderline predictor (adjusted $R^2=0.069$; $P= 0.038$) of mental health score. |
| Lin 2007(37)<br>SARS<br>Taiwan | Hospital<br>Physicians and nurses | N=92 | PTSD (DTS-C) | 86 of 92 (93.5%) medical staff considered the SARS outbreak to be a traumatic experience. The DTS-C scores of staff in the emergency department and in the psychiatric ward were significantly different ($P=0.04$). Emergency department staff had more severe PTSD symptoms than staff in the psychiatric ward |
| Marjanovic 2007(38)<br>SARS<br>Canada | Healthcare facilities<br>Nurses | N=333 | Emotional exhaustion (MBI-GS); state anger (adapted version of STAXI-2); Avoidance behavior (6 items) | Emotional exhaustion: Less contact with SARS patients ($\beta=-0.15$; $P=0.003$), higher levels of vigor ($\beta=0.34$; $P<0.001$), greater trust in equipment/infection control initiatives ($\beta=-0.15$; $P=0.005$), predicted significantly lower levels of emotional exhaustion.<br>Avoidance behavior: Less time spent in quarantine ($\beta = 0.23$; $P<0.001$), higher levels of vigor ($\beta=0.27$; $P<0.001$) and higher organizational support ($\beta=0.26$; $P<0.001$) predicted significantly less avoidance behavior.<br>State anger: Less time in quarantine ($\beta=0.14$; $P=0.008$), higher levels of organizational support ($\beta=0.24$; $P<0.001$) vigor ($\beta=-0.20$; $P<0.001$) and trust in equipment/infection control initiatives ($\beta=-0.14$; $P=0.011$) predicted lower levels of state anger |
| Wong 2007(39)<br>SARS<br>China and<br>Canada | Community<br>based primary<br>care clinics<br>Family<br>medicine<br>tutors | N=188 | Anxiety (a visual analogue<br>scale ranging from<br>extremely frightened<br>(high score) to not<br>frightened at all (zero<br>score). | 51% in Canada and 50.7% in Hong Kong, were classified in the high-anxiety<br>group. |
| Fiksenbaum<br>2006(40)<br>SARS<br>Canada | Multiple<br>settings<br>(community<br>health,<br>surgical and<br>pediatrics)<br>Nurses | N=333 | Emotional exhaustion<br>(MBI-GS); State anger<br>(STAXI) | Working conditions contributed significantly to an increase in perceived<br>SARS threat, which led to increased emotional exhaustion ($\beta=0.44$) and<br>state anger ($\beta=0.41$). Lower perceived organizational support was related<br>to increased feelings of emotional exhaustion ($\beta=0.11$) and state anger<br>($\beta=0.26$). |
| Lu 2006(41)<br>SARS<br>Taiwan | Hospital<br>HCWs:<br>physicians,<br>nurses and<br>others | N=127 | Extraversion, neuroticism (EPQ). Psychiatric Morbidity (CHQ). Paternal/maternal care and protection (PBI) | Twenty-two (17.3%) developed significant mental symptoms, and 105 (82.7%) showed no obvious symptoms. The structural equation model showed that maternal care ($\beta=-0.18$, $P = 0.011$) and neuroticism ($\beta=0.54$, $P<10^{-6}$) directly influenced the ability of HCWs to deal with the impact of SARS. Maternal overprotection ($\beta=-0.30$, $P< 0.05$) had an indirect influence on the ability to cope with the impact of SARS. |
| Maunder 2006(42)<br>SARS<br>Canada | Hospital<br>HCWs | N=769 | PTSD (IES) | From 13 to 26 months after the SARS outbreak, 769 HCWs at 9 Toronto hospitals that treated SARS patients and 4 Hamilton hospitals that did not treat SARS patients completed a survey of several adverse outcomes. Toronto HCWs reported significantly higher levels of burnout ($P=0.019$), psychological distress ($P<0.001$), and posttraumatic stress ($P<0.001$). Toronto workers were more likely to have reduced patient contact and work hours and to report behavioral consequences of stress. Variance in adverse outcomes was explained by a protective effect of the perceived adequacy of training and support and by a provocative effect of maladaptive coping style and other individual factors. |
| Ho 2005 (43)<br>SARS<br>China | Hospitals<br>HCWs | N=97 | PTSD (CIES-R) | Participants with lower self-efficacy tended to have higher fear related to SARS ($P<0.05$). The mean scores of the three CIES-R subscales ranged from 1.24 to 1.57, with Intrusion scoring highest. Fear related to SARS was also correlated positively with posttraumatic stress symptoms among respondents ($P<0.01$). |
| Phua 2005(44) | Hospital | N=99 | Psychiatric morbidity (IES and GHQ) | Psychiatric morbidity was 17.7% on the IES and 18.8% on the GHQ 28, with the trend for physicians to report lower psychiatric morbidity. |
| SARS<br>Singapore | Physicians and nurses |  |  |  |
| Tham 2005(45) | Hospital | N=96 | Depression, anxiety (GHQ 28), PTSD (IES), somatic symptoms, insomnia, social dysfunction (GHQ 28) | Thirty-eight out of 41 (92.7%) doctors and 58 out of 83 (69.9%) nurses responded. Fewer doctors reported post-event and psychiatric morbidity compared to nurses, with 5 (13.2%) doctors and 12 (20.7%) nurses scoring $\geq 26$ on IES, 6 (15.8%) doctors and 12 (20.7%) nurses scoring $\geq 5$ on GHQ 28. The doctors reported a median of 9.5 (range 0-47) on IES and 0 (range 0-11) on GHQ 28. The nurses reported a median of 15 (range 0-61) on IES and 1 (range 0-25) on GHQ 28. |
| SARS<br>Singapore | Physicians and nurses |  |  |  |
| Wong 2005(46) | Emergency Department of public hospital | N=466 | Mental distress (unclear), coping strategies (Chinese version of the Brief Cope) | The mean overall distress level was 6.19 out of a 10-point scale. |
| SARS<br>China | HCWs |  |  |  |
| Chua 2004 (47) | Hospitals | N=613 | StressPSS-10 | Stress levels were raised in both groups (PSS=18) but were not relatively increased in the HCWs. HCWs reported significantly more positive (94%, N=256) and more negative psychological effects (89%, N=241) from SARS than did control subjects. HCWs declared confidence in infection-control measures. |
| SARS<br>China | HCWs, mostly nurses (60%) |  |  |  |
| Tam 2004(48)<br>SARS<br>China | Hospital<br>(medical units<br>and intensive<br>care units)<br>HCWs | N=652 | Subjective job related<br>stress levels (2 Likert-type<br>questions); Psychological<br>morbidity (GHQ-12) | 68% reported a high level of stress. About 57% were found to have<br>experienced psychological distress. The HCWs' psychological morbidity was<br>best understood by the perceptions of personal vulnerability, stress and<br>support in the workplace. |
| Verma 2004(49)<br>SARS<br>Singapore | Unspecified<br>Physicians and<br>Traditional<br>Chinese<br>Medicine<br>(TCM)<br>Practitioners | N=1050 | Depression, anxiety (GHQ<br>28), PTSD (IES),<br>perception of stigma<br>(Questionnaire adopted<br>from the HSS). | A total of 721 (29%) GPs and 329 (22%) TCM practitioners responded to<br>the survey. Significantly more GPs had worked in SARS affected facilities<br>and had been directly involved in the care of patients with SARS than the<br>TCM practitioners ($P<0.001$). Those GPs who were directly involved in the<br>care of patients with SARS were significantly more likely to be GHQ cases<br>as compared to those not involved in the care of patients with SARS<br>($P=0.02$; $OR=2.9$; 95% CI, 1.3 to 6.3). The mean score of the GHQ somatic,<br>anxiety and social dysfunction subscales were significantly higher in GPs as<br>compared to TCM Practitioners ($P<0.001$). The GHQ total score as well as<br>the subscales was significantly correlated with the IES-R and stigma<br>subscales ($P<0.05$). |
| Wong 2004(50)<br>SARS<br>China | Community<br>based primary<br>care clinics<br>Medicine<br>Tutors | N=137 | Anxiety (unclear) | All agreed SARS had changed their clinical practices. Significant anxiety was<br>found in family doctors. Three quarters of respondents recalled requesting<br>more investigations while a quarter believed they had over-prescribed<br>antibiotics. GPs who were exposed to SARS or who had worked in high<br>infection districts were less likely to quarantine themselves (10.8% versus<br>33.3%; $P<0.01$; 6.5% versus 27.5%; $P<0.01$ respectively). Exposure to SARS,<br>the infection rates in their working district, and anxiety levels had<br>significant impact on the level of protection or prescribing behaviour. |

| **Systematic review** |  |  |  |  |
| --- | --- | --- | --- | --- |
| Brooks 2018<br>(51) | Multiple<br>settings | NR (22 studies) | Depression, anxiety,<br>PTSD, job-related stress | The psychological impact of SARS on HCWs appeared to be associated with occupational role; training/preparedness; high-risk work environments; quarantine; role-related stressors; perceived risk; social support; social rejection/isolation; and impact of SARS on personal or professional life. |
| SARS | HCWs |  |  |  |
| Worldwide |  |  |  |  |
| **Cohort (prospective) studies** |  |  |  |  |
| Lung 2009(52) | Hospital | N=127 | Extraversion, neuroticism<br>(EPQ). Psychiatric<br>Morbidity (CHQ).<br>Paternal/maternal care<br>and protection (PBI) | The first stage symptom results had a positive direct effect on the second stage results (somatic symptoms: $\beta=0.31$, $P<0.05$; negative affect: $\beta=0.23$, $P=0.006$). The participants presented more somatic symptoms than negative affect at both stages (accounting for 35% and 19% of the variance, respectively). Daily-life stressful events had impact on both dimensions of mental health, which included somatic symptoms ($\beta=0.18$, $P=0.034$) and negative affect ($\beta=0.30$, $P<0.05$). Nurses had fewer somatic realm than other healthcare workers ($\beta=-0.15$, $P<0.05$). Participants who had negative effect at the initial assessment had fewer somatic symptoms ($\beta=-0.21$, $P<0.05$) the following year. Negative affect had impact on somatic symptoms at both assessment stages (initial stage: $\beta=0.25$, $P<0.05$; follow-up stage: $\beta=0.24$, $P<0.05$) |
| SARS | HCWs:<br>doctors,<br>nurses and<br>others<br>(medical<br>technicians,<br>respiratory<br>therapists, or<br>emergency<br>attendant) |  |  |  |
| Taiwan |  |  |  |  |
| Wu 2009(53)<br>SARS<br>China | Hospitals<br>HCWs | N=549 | PTSD (IES-R) | About 10% of the respondents had experienced high levels of posttraumatic stress (PTS) symptoms since the SARS outbreak. Respondents who had been quarantined, or worked in high-risk locations such as SARS wards, or had friends or close relatives who contracted SARS, were 2 to 3 times more likely to have high PTS symptom levels, than those without these exposures. Respondents' perceptions of SARS-related risks were significantly positively associated with PTS symptom levels and partially mediated the effects of exposure. Altruistic acceptance of work-related risks was negatively related to PTS levels. |
| McAlonan 2007(54)<br>SARS<br>China | Hospitals<br>HCWs | N=176 | Depression, anxiety (DASS-21), PTSD (IES), stress (PSS-10) | In 2003, high-risk health care workers had elevated stress levels (PSS-10 score = 17.0) that were not significantly different from levels in low-risk HCW control subjects (PSS-10 score=15.9). More high-risk HCWs reported fatigue, poor sleep, worry about health, and fear of social contact, despite their confidence in infection-control measures. By 2004, however, stress levels in the high-risk group were not only higher (PSS-10 score=18.6) but also significantly higher than scores among low-risk HCW control subjects (PSS-10 score=14.8, $P<0.05$). In 2004, the perceived stress levels in the high-risk group were associated with higher depression, anxiety, and posttraumatic stress scores ($P<0.001$). |

CES-D= Center for Epidemiologic Studies Depression Scale; CHQ= Chinese Health Questionnaire; CI= confidence interval; CIES-R= Chinese Impact of Event Scale—Revised; DASS-21: Depression= Anxiety and Stress Scale – 21 Items; DRS-15= Dispositional Resilience Scale-15; DTS-C= Davidson Trauma Scale Chinese Version; EPQ= Eysenck Personality Questionnaire; GAD-7= Generalized Anxiety Disorder 7-item; GHQ= General Health Questionnaire; GP= general
practitioner; H1N1: H1N1 influenza virus; HCWs= health care workers; HrQoL= Health-related quality of life; HSS= HIV Stigma Scale; IES= Impact of Events Scales; IES-R= Impact of Event Scale-Revised; MBI-GS= Maslach Burnout Inventory —General Survey; MERS-CoV: Middle East respiratory syndrome coronavirus; MOS SF-36= Medical Outcome Study Short-Form 36 Survey; OR= odds ratio; PBI= Parental Bonding Instrument; PCL-C= PTSD Checklist-Civilian Version; PHQ-9= Patient Health Questionnaire-9; PSS-10= 10-item Perceived Stress Scale; PTSD= Post-Traumatic Stress Disorder; SARS= severe acute respiratory syndrome; SE= standard error; SF12= 12-item Short Form Health Survey; SF-36= Short Form-36 questionnaire; STAXI-2= State-Trait Anger Expression Inventory.

**C. Summary of Findings table of studies examining the impact on mental health problems in healthcare workers during and after viral epidemics (N= 2)**

| **Author/**<br>**year/health**<br>**emergency/**<br>**country/** | **Setting**<br>**Type of**<br>**healthcare**<br>**professionals** | **Sample size (N)** | **Mental health problem**<br>**(tool)** | **Main results** |
| --- | --- | --- | --- | --- |
| Lee 2007(55)<br>SARS<br>China | Hospitals<br>HCWs | Cohort<br>(prospective)<br>N=33 | Depression, anxiety (DASS-21), Subjective stress (PSS-10), PTSD (IER-R), psychiatric morbidity (GHQ-12) | SARS survivors had higher stress levels during the outbreak, compared with control subjects (PSS-10 scores=19.8 and 17.9, respectively; $P<0.01$). SARS survivors had stress levels similar to those of non-HCWs, but HCWs showed significantly higher stress levels in 2004 (PSS-10 score=22.8, compared with PSS-10 score=18.4; $P<0.05$) and had higher depression ($P<0.01$), anxiety ($P<0.001$), posttraumatic symptoms (Intrusion $P<0.001$, Avoidance $P<0.05$ and Hyperarousal $P<0.05$), and GHQ-12 scores ($P<0.001$). |
| Chong 2004(56)<br>SARS<br>China | Tertiary hospital<br>HCWs | Cross-sectional<br>N=1,257 | Depression (CHQ), anxiety (CHQ), intrusion and avoidance (IES), and somatization (CHQ) | In the initial phase of the outbreak, when the infection was spreading rapidly, feelings of extreme vulnerability, uncertainty and threat to life were perceived, dominated by somatic and cognitive symptoms of anxiety. During the 'repair' phase, when the infection was being brought under control, depression and avoidance were evident. Prevalence of psychiatric morbidity (CHQ) about 75%. |

CHQ= Chinese Health Questionnaire; DASS-21= Depression= Anxiety and Stress Scale – 21 Items; GHQ-12= 12-item General Health Questionnaire; HCWs= healthcare workers; IES= Impact of Events Scales; PSS-10= Perceived Stress Scale; PTSD= Post-Traumatic Stress Disorder; SARS= severe acute respiratory syndrome.

## Online Appendix 5 Forest plots

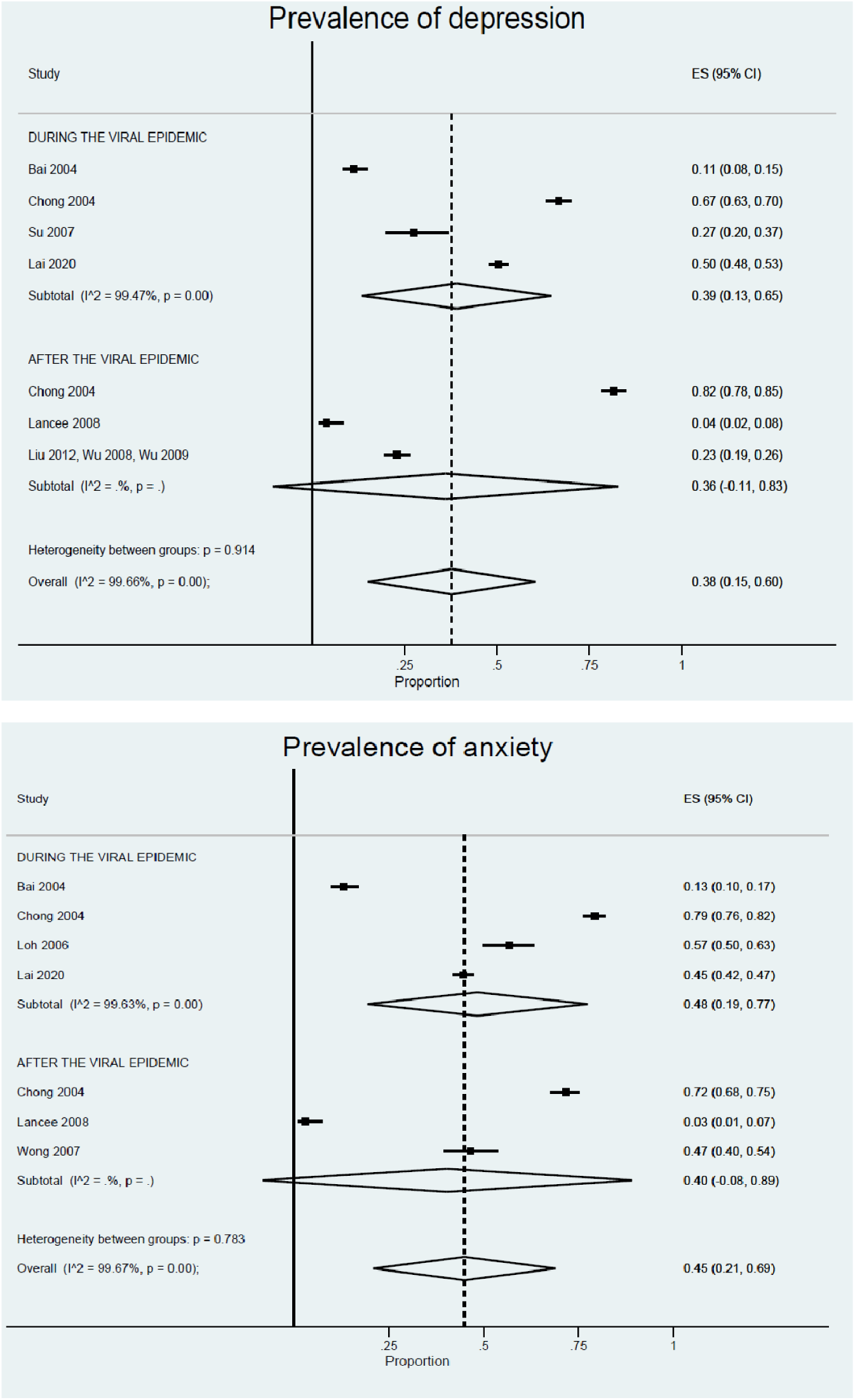
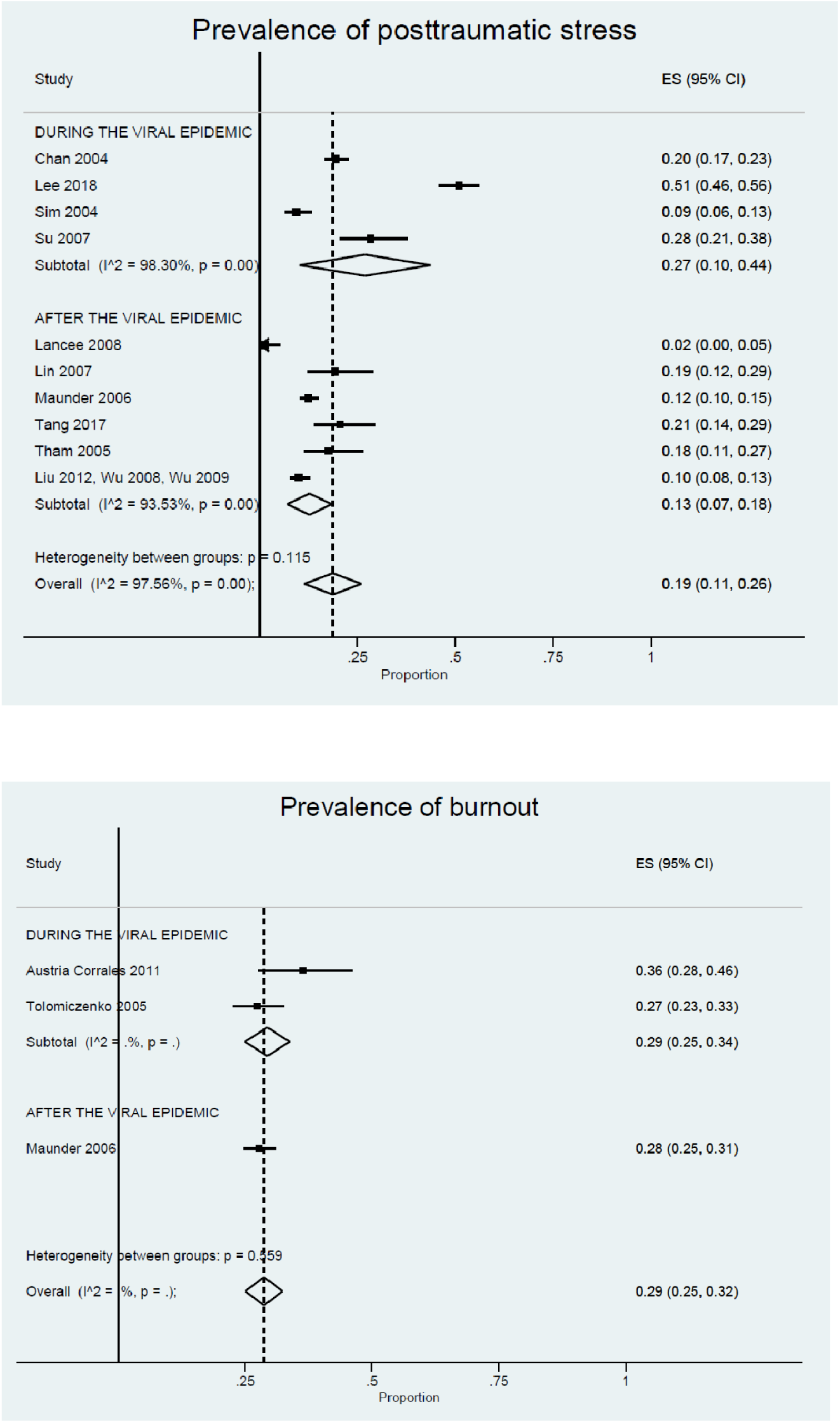
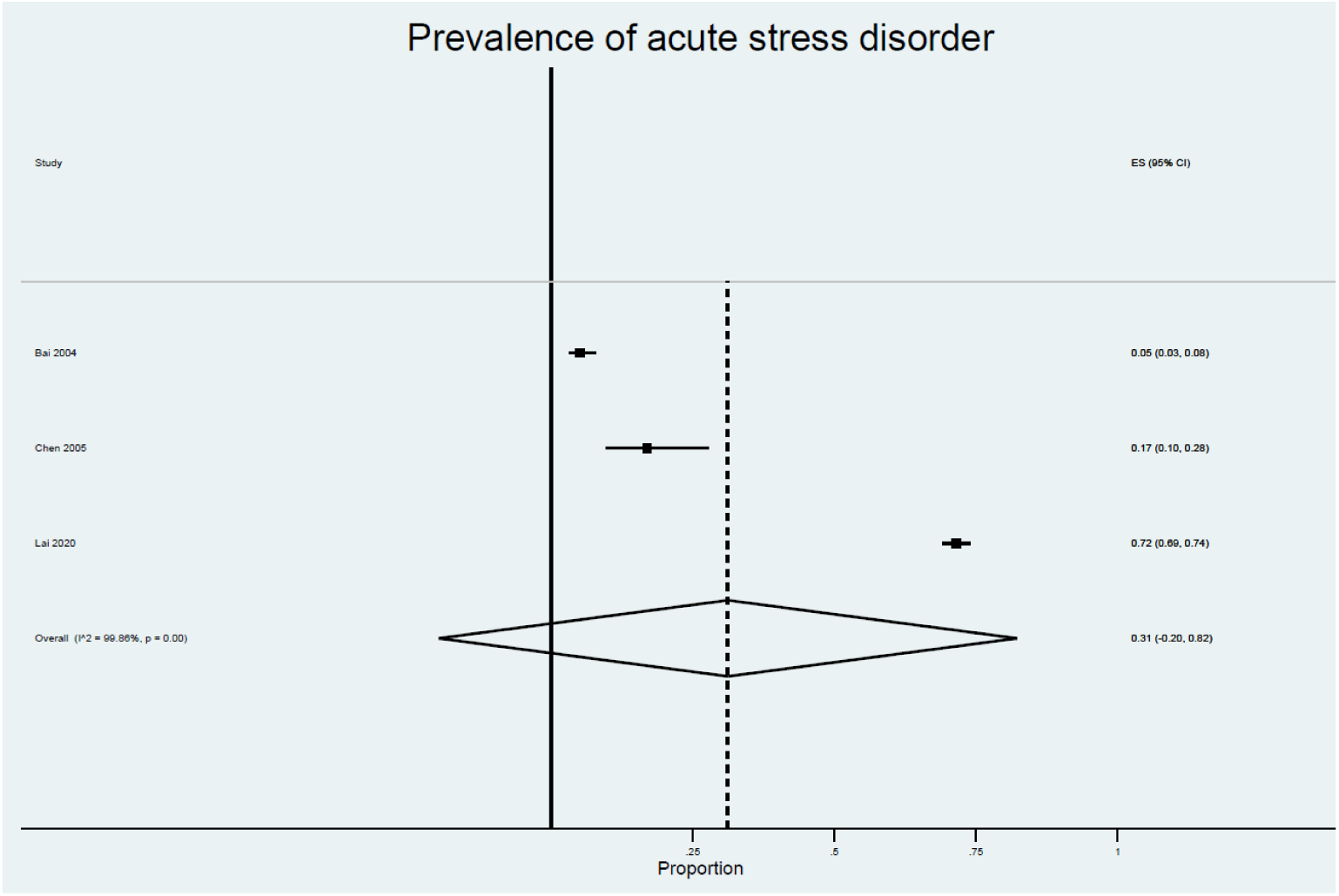

## Online Appendix 6 Summary of Findings table of studies of interventions to reduce impact of epidemic outbreaks on mental health of healthcare professionals

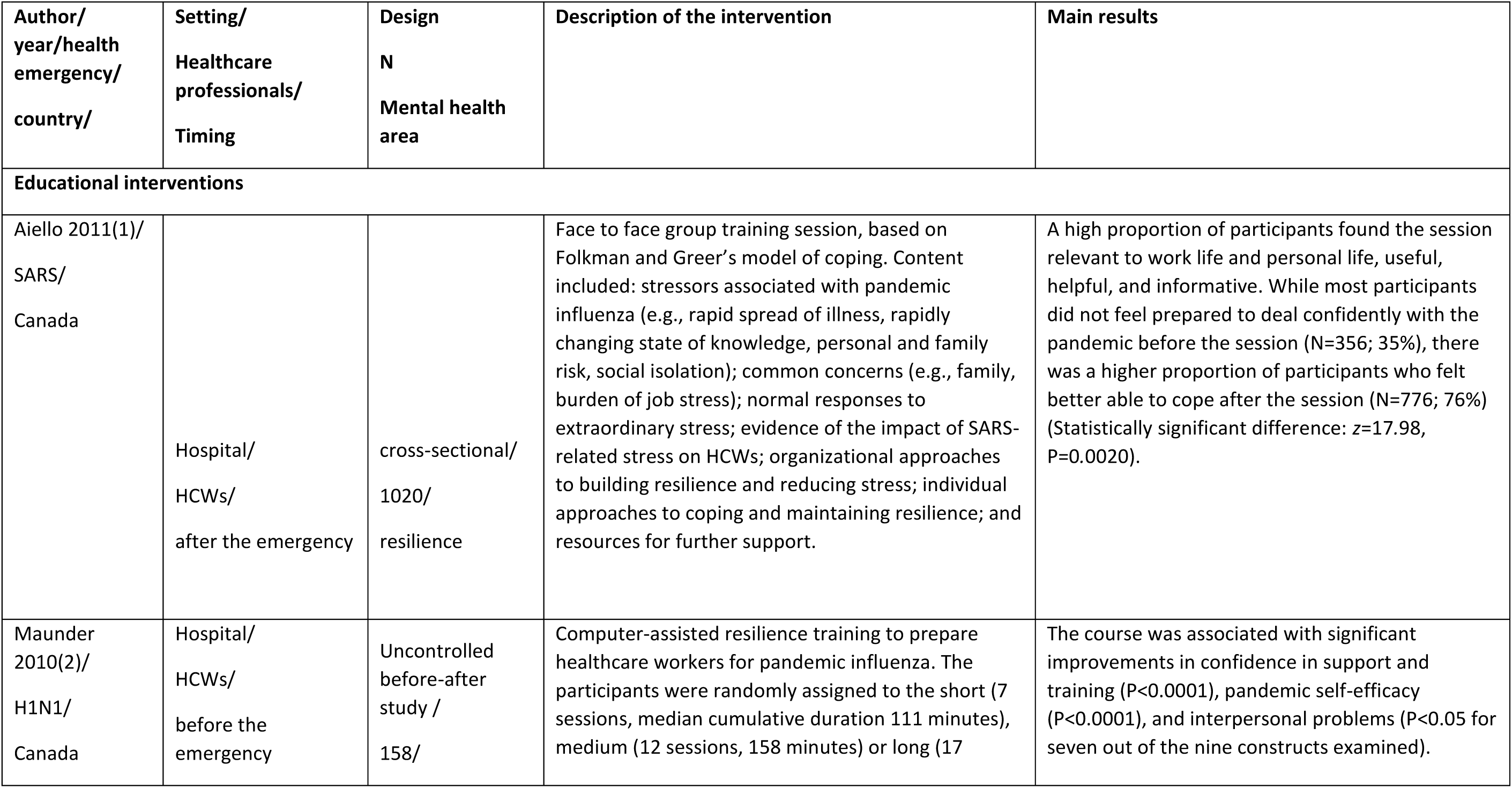
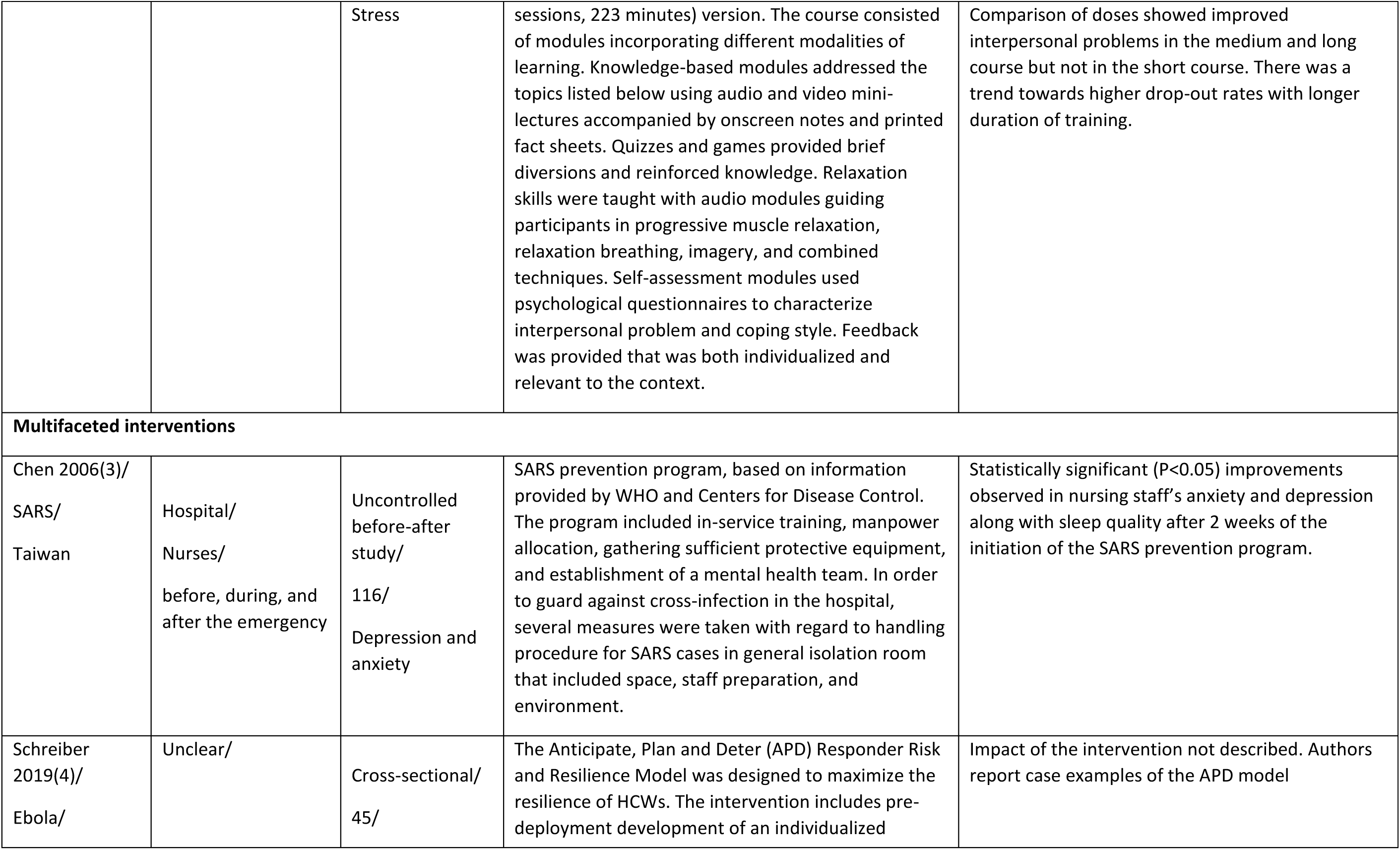
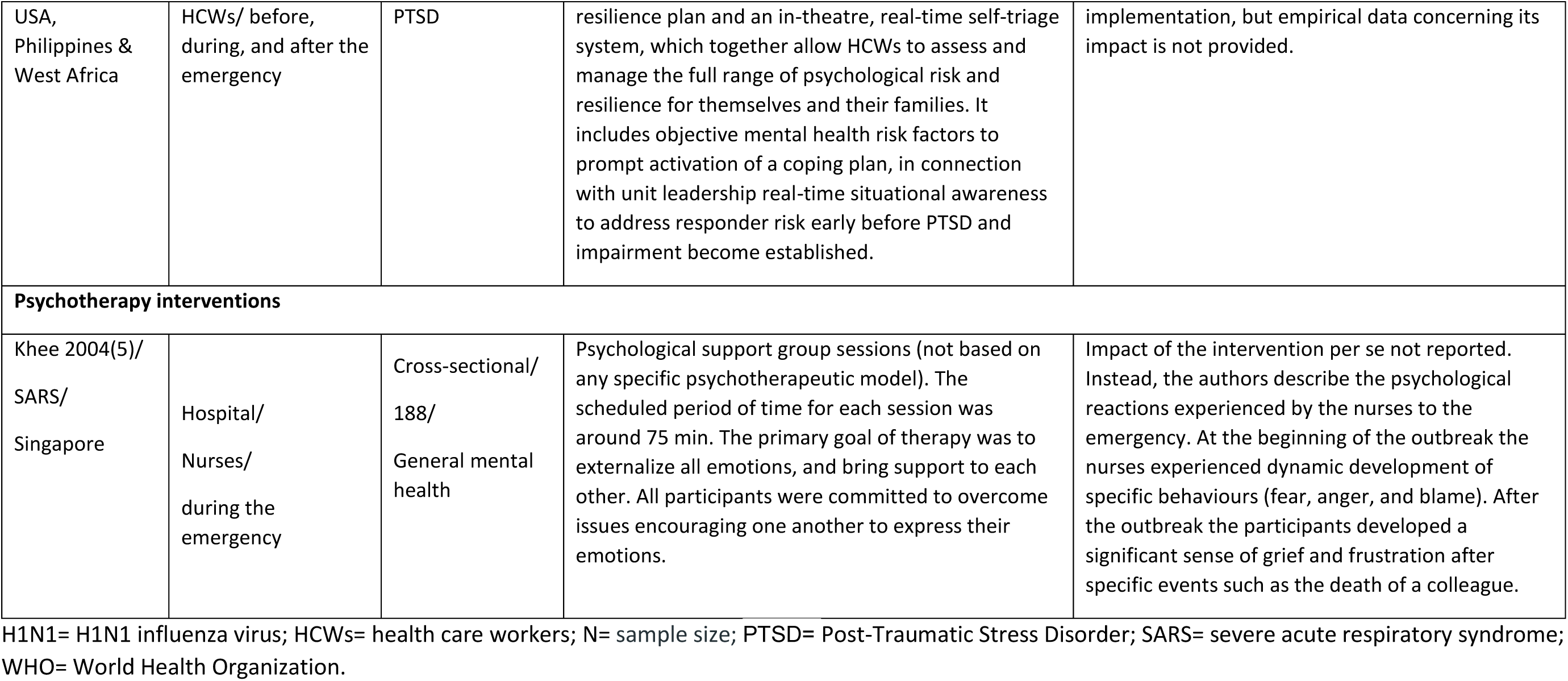

| Author/<br>year/health<br>emergency/<br>country/ | Setting/<br>Healthcare<br>professionals/<br>Timing | Design<br>N<br>Mental health<br>area | Description of the intervention | Main results |
| --- | --- | --- | --- | --- |
| **Educational interventions** |  |  |  |  |
| Aiello 2011(1)/<br>SARS/<br>Canada | Hospital/<br>HCWs/<br>after the emergency | cross-sectional/<br>1020/<br>resilience | Face to face group training session, based on Folkman and Greer's model of coping. Content included: stressors associated with pandemic influenza (e.g., rapid spread of illness, rapidly changing state of knowledge, personal and family risk, social isolation); common concerns (e.g., family, burden of job stress); normal responses to extraordinary stress; evidence of the impact of SARS-related stress on HCWs; organizational approaches to building resilience and reducing stress; individual approaches to coping and maintaining resilience; and resources for further support. | A high proportion of participants found the session relevant to work life and personal life, useful, helpful, and informative. While most participants did not feel prepared to deal confidently with the pandemic before the session (N=356; 35%), there was a higher proportion of participants who felt better able to cope after the session (N=776; 76%) (Statistically significant difference: $z=17.98$, $P=0.0020$). |
| Maunder 2010(2)/<br>H1N1/<br>Canada | Hospital/<br>HCWs/<br>before the<br>emergency | Uncontrolled<br>before-after<br>study /<br>158/ | Computer-assisted resilience training to prepare healthcare workers for pandemic influenza. The participants were randomly assigned to the short (7 sessions, median cumulative duration 111 minutes), medium (12 sessions, 158 minutes) or long (17 | The course was associated with significant improvements in confidence in support and training ($P<0.0001$), pandemic self-efficacy ($P<0.0001$), and interpersonal problems ($P<0.05$ for seven out of the nine constructs examined). |
|  |  | Stress | sessions, 223 minutes) version. The course consisted of modules incorporating different modalities of learning. Knowledge-based modules addressed the topics listed below using audio and video mini-lectures accompanied by onscreen notes and printed fact sheets. Quizzes and games provided brief diversions and reinforced knowledge. Relaxation skills were taught with audio modules guiding participants in progressive muscle relaxation, relaxation breathing, imagery, and combined techniques. Self-assessment modules used psychological questionnaires to characterize interpersonal problem and coping style. Feedback was provided that was both individualized and relevant to the context. | Comparison of doses showed improved interpersonal problems in the medium and long course but not in the short course. There was a trend towards higher drop-out rates with longer duration of training. |
| **Multifaceted interventions** |  |  |  |  |
| Chen 2006(3)/<br>SARS/<br>Taiwan | Hospital/<br>Nurses/<br>before, during, and<br>after the emergency | Uncontrolled<br>before-after<br>study/<br>116/<br>Depression and<br>anxiety | SARS prevention program, based on information provided by WHO and Centers for Disease Control. The program included in-service training, manpower allocation, gathering sufficient protective equipment, and establishment of a mental health team. In order to guard against cross-infection in the hospital, several measures were taken with regard to handling procedure for SARS cases in general isolation room that included space, staff preparation, and environment. | Statistically significant ($P<0.05$) improvements observed in nursing staff's anxiety and depression along with sleep quality after 2 weeks of the initiation of the SARS prevention program. |
| Schreiber<br>2019(4)/<br>Ebola/ | Unclear/ | Cross-sectional/<br>45/ | The Anticipate, Plan and Deter (APD) Responder Risk and Resilience Model was designed to maximize the resilience of HCWs. The intervention includes pre-deployment development of an individualized | Impact of the intervention not described. Authors report case examples of the APD model |
| USA, Philippines & West Africa | HCWs/ before, during, and after the emergency | PTSD | resilience plan and an in-theatre, real-time self-triage system, which together allow HCWs to assess and manage the full range of psychological risk and resilience for themselves and their families. It includes objective mental health risk factors to prompt activation of a coping plan, in connection with unit leadership real-time situational awareness to address responder risk early before PTSD and impairment become established. | implementation, but empirical data concerning its impact is not provided. |
| **Psychotherapy interventions** |  |  |  |  |
| Khee 2004(5)/ SARS/ Singapore | Hospital/ Nurses/ during the emergency | Cross-sectional/ 188/ General mental health | Psychological support group sessions (not based on any specific psychotherapeutic model). The scheduled period of time for each session was around 75 min. The primary goal of therapy was to externalize all emotions, and bring support to each other. All participants were committed to overcome issues encouraging one another to express their emotions. | Impact of the intervention per se not reported. Instead, the authors describe the psychological reactions experienced by the nurses to the emergency. At the beginning of the outbreak the nurses experienced dynamic development of specific behaviours (fear, anger, and blame). After the outbreak the participants developed a significant sense of grief and frustration after specific events such as the death of a colleague. |

H1N1= H1N1 influenza virus; HCWs= health care workers; N= sample size; PTSD= Post-Traumatic Stress Disorder; SARS= severe acute respiratory syndrome; WHO= World Health Organization.

## Online Appendix 7 Evidence profiles of the certainty of evidence concerning interventions for preventing the psychological impact of infectious epidemic outbreaks in healthcare workers

**Evidence profile 1: Educational interventions compared to usual care for preventing the psychological impact of infectious epidemic outbreaks in healthcare workers**

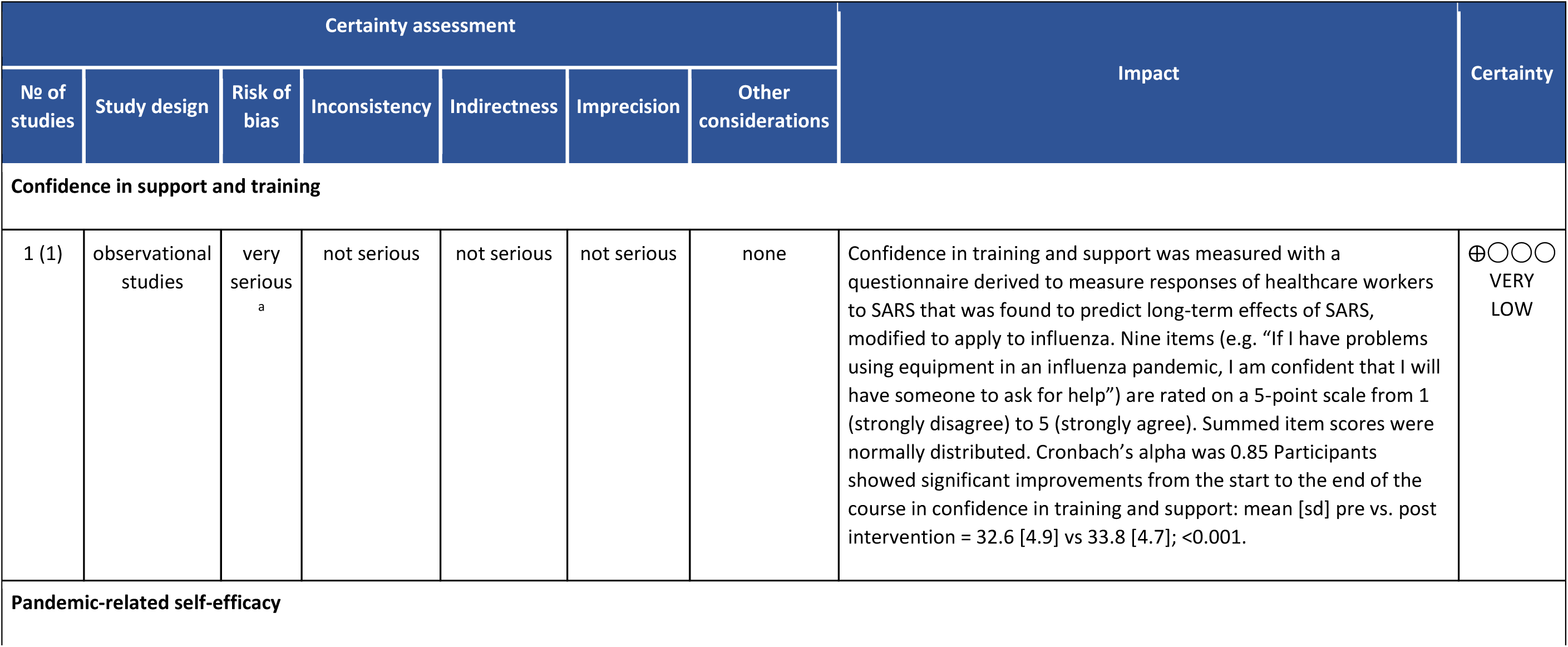
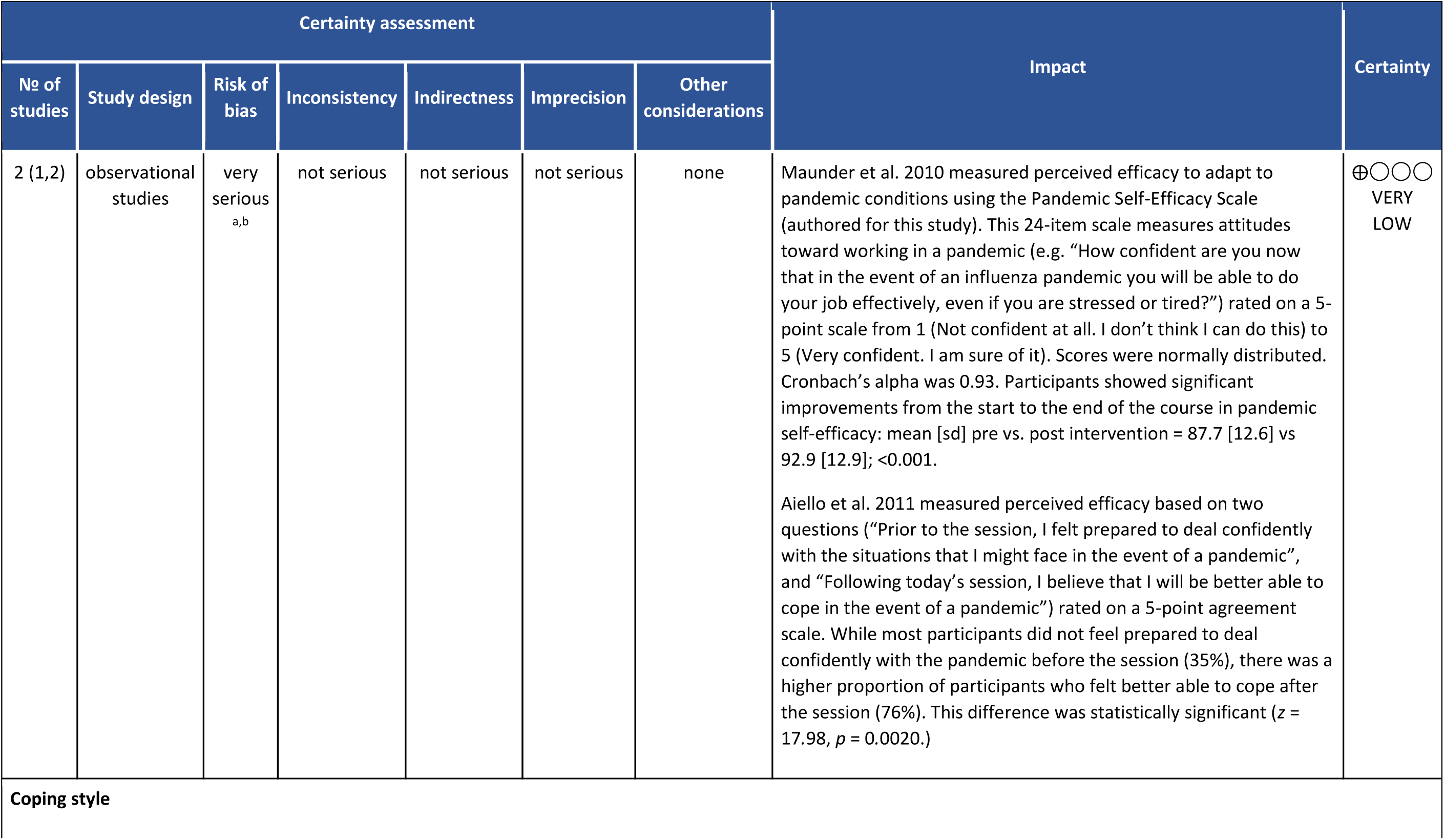
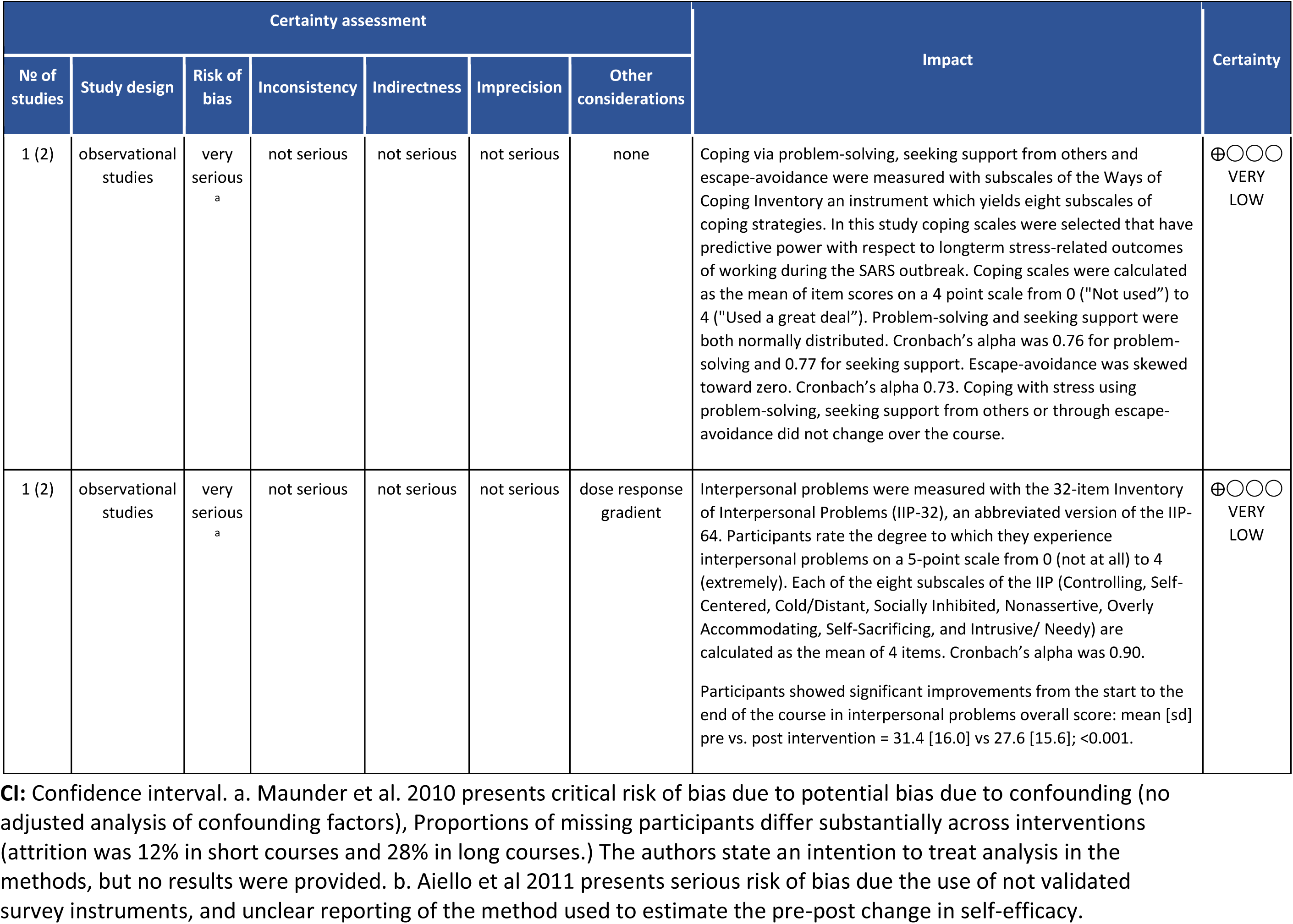

**Evidence profile 2. Multifaceted interventions compared to usual care for reducing the psychological impact during infectious epidemic outbreaks in healthcare workers**

| Certainty assessment |  |  |  |  |  |  | Impact | Certainty |
| --- | --- | --- | --- | --- | --- | --- | --- | --- |
| No of studies | Study design | Risk of bias | Inconsistency | Indirectness | Imprecision | Other considerations |  |  |
| **Anxiety** |  |  |  |  |  |  |  |  |
| 1 (3) | observational studies | serious<sup>a</sup> | not serious | not serious | not serious | none | Anxiety was measured with Zung's self-rating anxiety scale (SAS). After implementation of the SARS prevention program, the mean scores of anxiety level decreased with time. The mean score from the first questionnaire before caring for SARS patients is 60 points (SD= 9.28), indicating moderate anxiety compared to the mean scores of after the implementation of the anti-SARS program, the mean scores of 51, 50, and 46 indicating mild anxiety, mild anxiety, no anxiety, respectively, at the remaining time points. GEE models showed that anxiety levels 2 weeks after the implementation of the prevention program and while caring for SARS patients were significantly lower than anxiety level before caring for SARS patients (p = 0:075). The anxiety level month after caring for SARS patients was also significantly lower than the anxiety levels before care of SARS patients (p<0:0001). After caring for SARS patients for 3 months and 1 month after the hospital returned to normal operations, nursing personnel exhibited anxiety levels significantly lower than that prior to caring for SARS patients (p<0:0001). | ⊕○○○<br>VERY LOW |
| **Depression** |  |  |  |  |  |  |  |  |
| No of studies | Study design | Risk of bias | Inconsistency | Indirectness | Imprecision | Other considerations |  |  |
| 1 (3) | observational studies | serious<sup>a</sup> | not serious | not serious | not serious | none | Depression was measured with Zung's self-rating depression scale (SDS). After the implementation of the SARS prevention program, mean scores of depression level decreased with time. At the time of the first questionnaire before care of SARS patients began, the mean score was 61 points (SD =12.62), indicating moderate depression. After program initiation, the mean scores of 51, 50, and 48 points indicated mild depression, mild depression, and no depression, respectively. Using GEE, the levels of depression 2 weeks after initiation of the SARS prevention program and while caring for SARS patients were significantly lower than the level before caring for SARS patients ($p<0:0001$). At 1 month after caring for SARS patients, the level of depression was also significantly lower than the levels of depression levels before the staff began to care for SARS patients ($p<0:0001$). After the hospital had returned to normal operations, the level of depression was significantly lower than that prior to taking care of SARS patients ($p<0:0001$). | ⊕○○○<br>VERY LOW |
| **Sleep quality** |  |  |  |  |  |  |  |  |
| 1 (3) | observational studies | serious<sup>a</sup> | not serious | not serious | not serious | none | Sleep quality was measured with the Pittsburgh sleep quality index. The mean score from the first questionnaire was 12 points (SD =3.83), meaning that sleep quality was poor. Of the remaining three questionnaires, the mean scores were 10, 10, and 8 indicating poor sleep quality. GEE showed that the mean score at 2 weeks after the SARS prevention program began when the staff was caring for SARS patients was significantly lower than the mean score before they began caring for SARS patients ($p=0.0053$). At 1 month after caring for SARS patients and at 1 month after the hospital resumed normal operations, the mean score ($p=0:0017$) was also significantly lower than the score before the nursing care began ($p=0:0008$). | ⊕○○○<br>VERY LOW |

CI: Confidence interval. a. Chen et al. 2006 presents serious risk of bias due to serious risk of selection bias (nurses volunteered to participate in the study - participation probably related to intervention/outcome), and serious bias in measurement of outcomes (subjective measures, and the team was aware of allocation status.)

## Online Appendix 8 PRISMA checklist

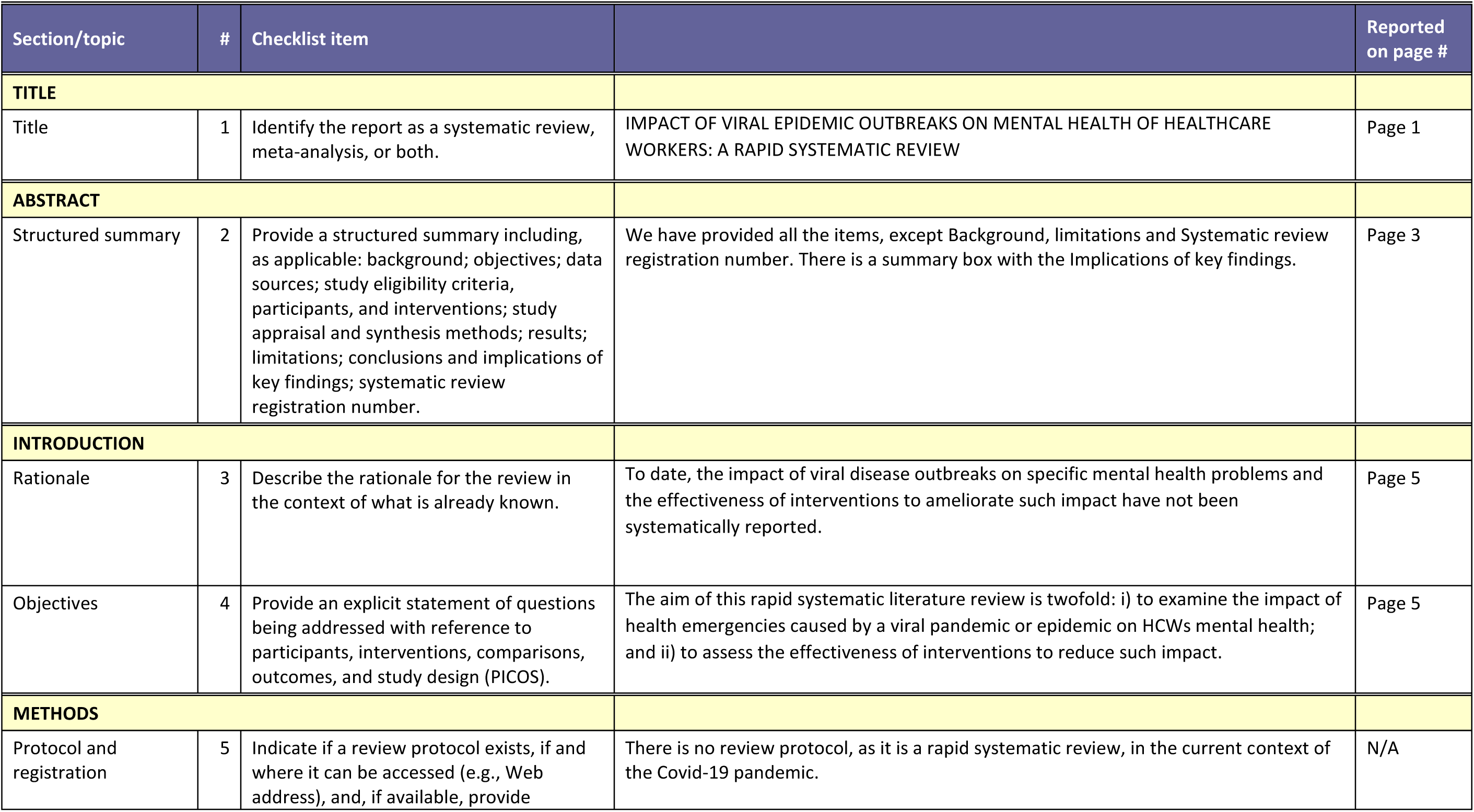
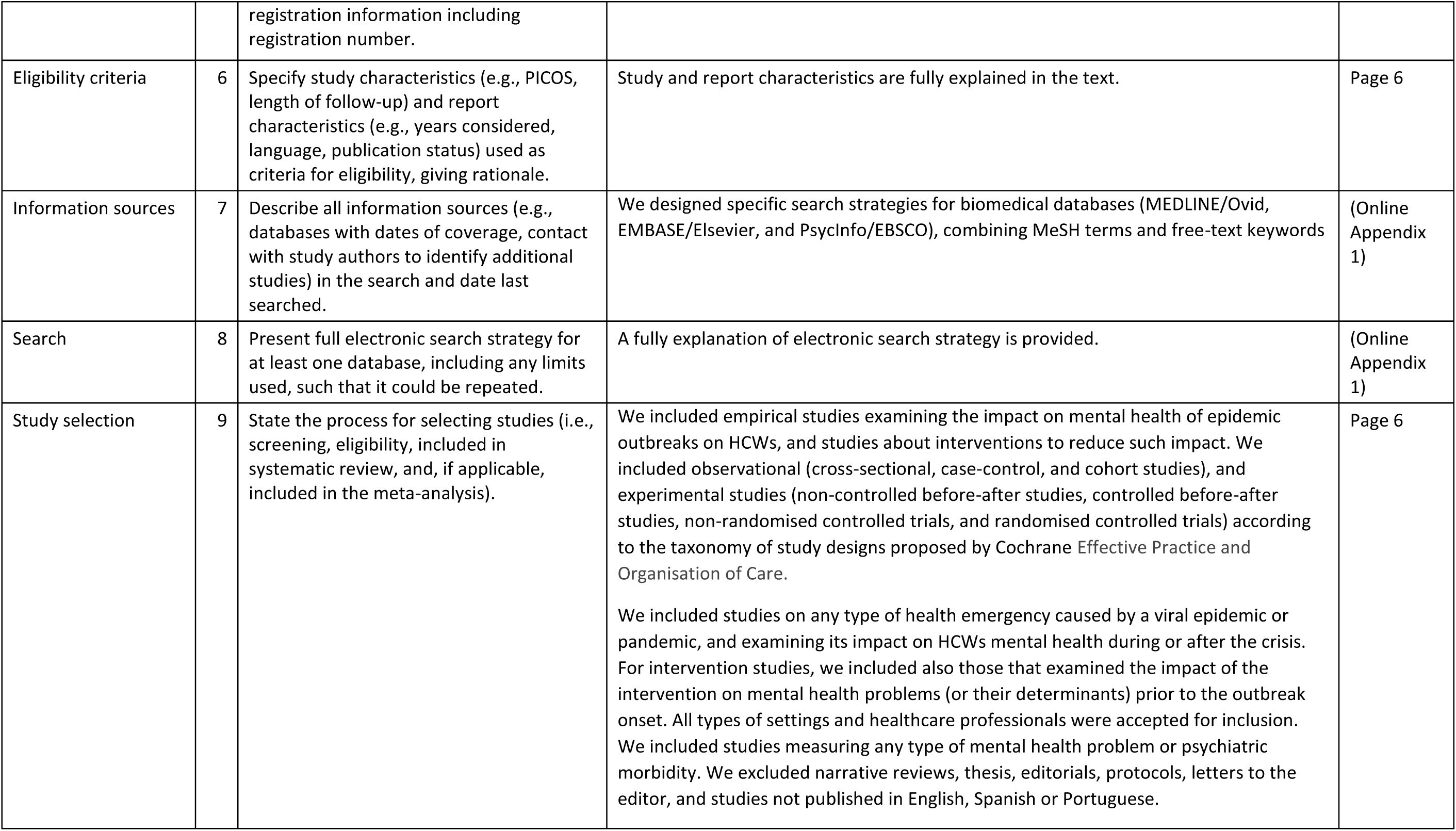
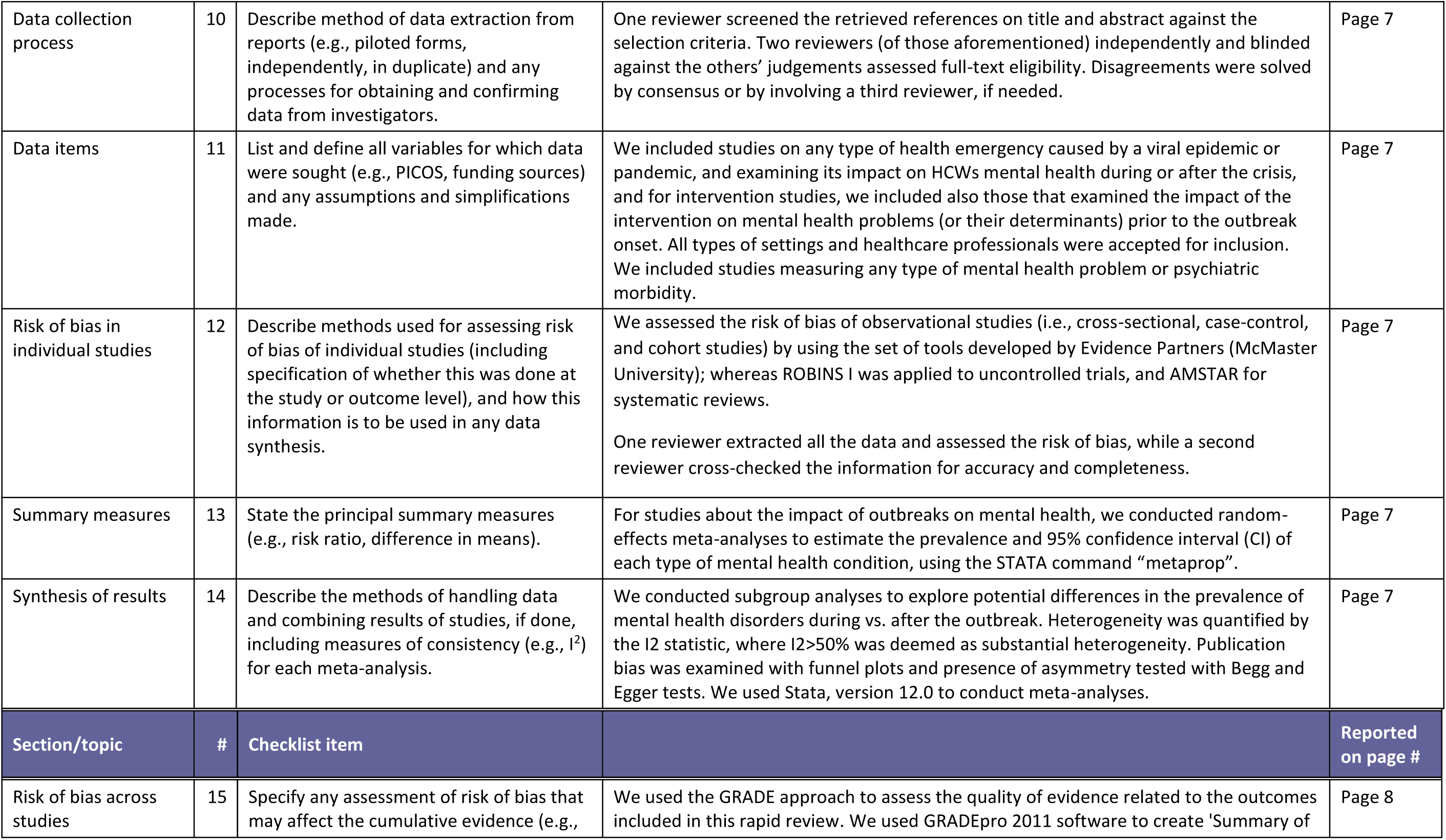
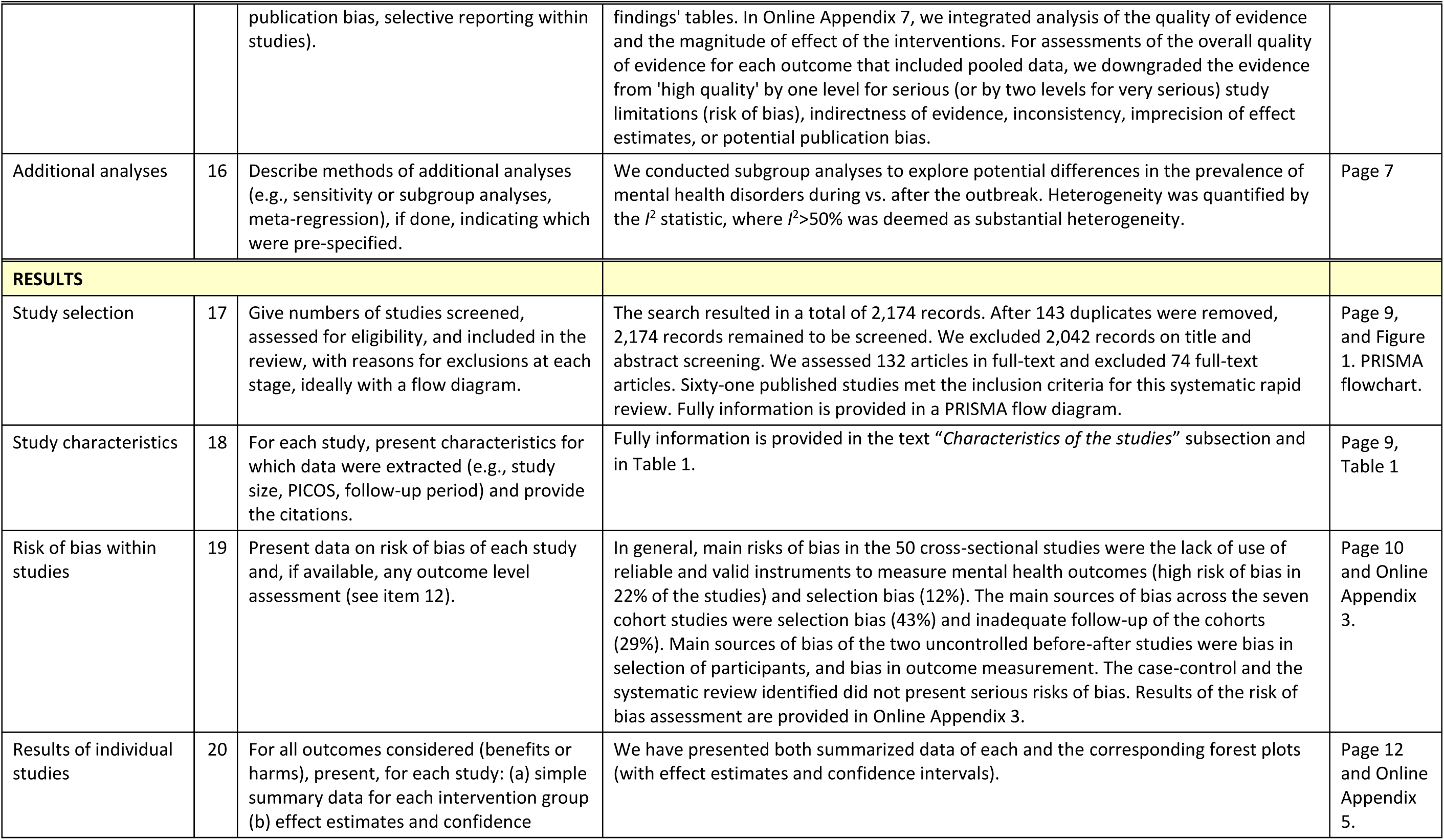
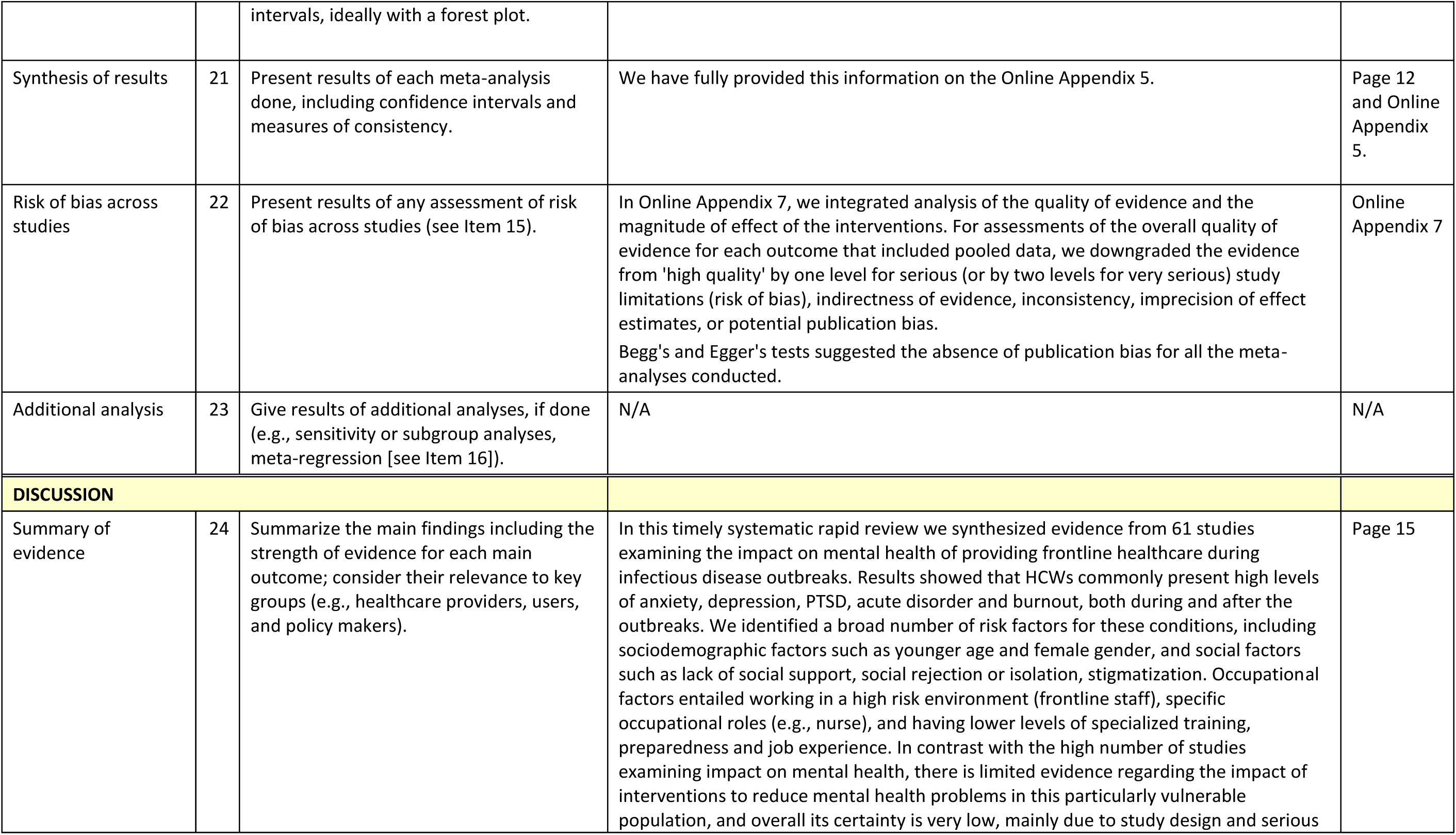
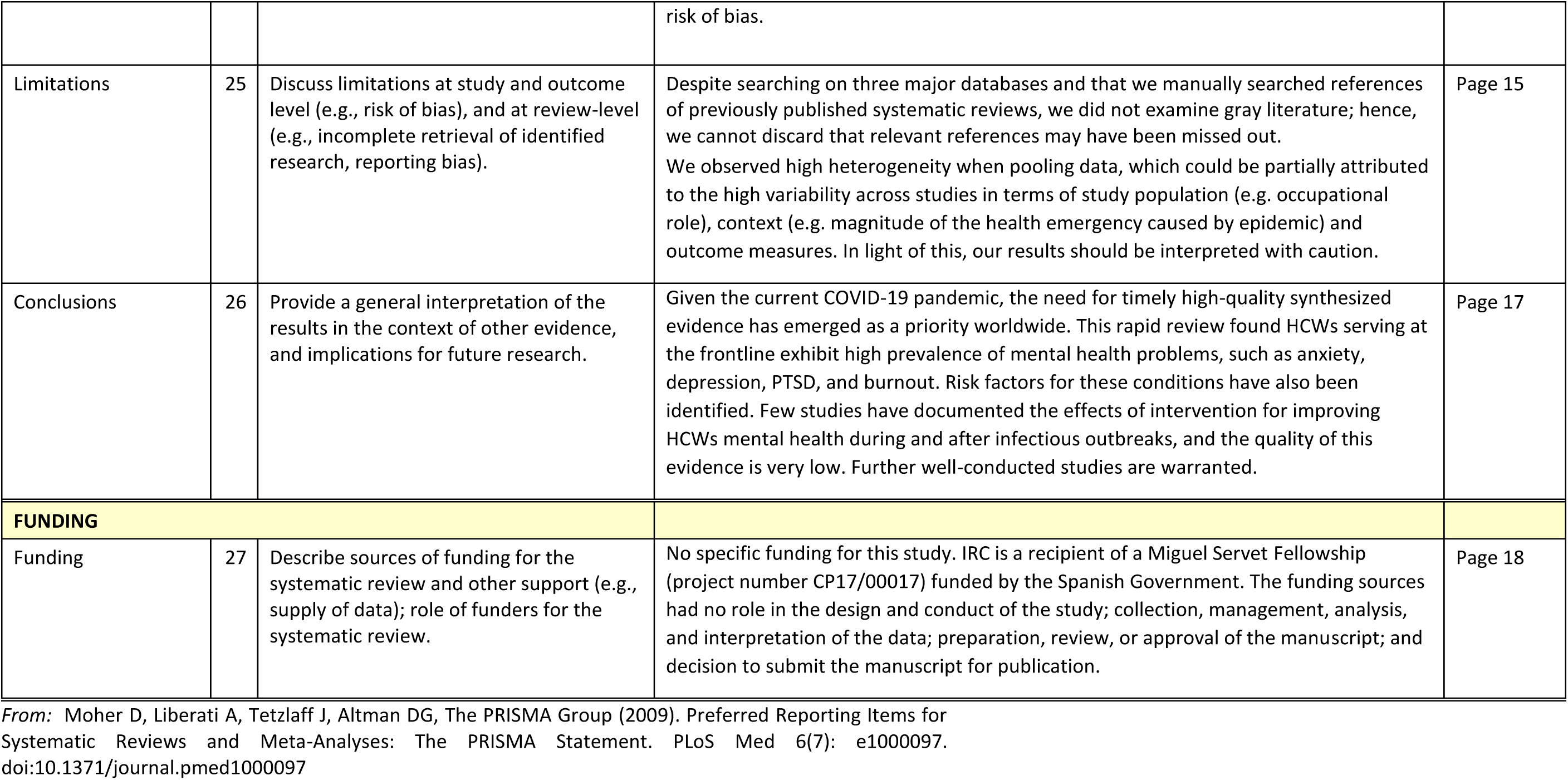

## Notes

### Competing Interest Statement

The authors have declared no competing interest.

